# Dose-response associations of intermittent lifestyle physical activity micropatterns and incident type 2 diabetes

**DOI:** 10.64898/2026.01.19.26344379

**Authors:** Kar Hau Chong, Matthew N. Ahmadi, Raaj K. Biswas, Monique E. Francois, Nicholas A. Koemel, Angelo Sabag, Martin J. Gibala, Shelley E. Keating, Jonathan Little, Cecilie Thøgersen-Ntoumani, Emmanuel Stamatakis

## Abstract

**Objective:** To examine dose-response associations of vigorous intermittent lifestyle physical activity micropatterns (VILPA; bouts ≤1 minute) and its moderate-to-vigorous equivalent (MV-ILPA; bouts ≤3 minutes) with incident type 2 diabetes.

**Research Design and Methods:** Prospective data from UK Biobank accelerometry sub-study participants who reported no leisure-time exercise and had no type 2 diabetes at baseline were analysed. Daily duration and frequency of VILPA and MV-ILPA were derived from wrist-worn accelerometry. Incident type 2 diabetes was ascertained through linked primary care, hospital and death records. Dose-response associations were examined using multivariable-adjusted Fine-Gray subdistribution hazard models accounting for competing risks.

**Results:** Among 22,706 participants (mean age 62.3 years; 56.4% female), 665 developed type 2 diabetes over an average follow-up of 7.9 years. Daily durations of VILPA and MV-ILPA showed inverse, non-linear L-shaped associations with incident type 2 diabetes. Median durations of 3.9 minutes/day of VILPA and 25.3 minutes/day of MV-ILPA were associated with 36% and 46% lower risk of type 2 diabetes, respectively, compared with those completing no VILPA or 3.9 minutes/day of MV-ILPA. Daily VILPA frequency showed a near-linear inverse association, with 10.4 bouts/day (median) corresponding to a hazard ratio (HR) of 0.64 (95% CI 0.51-0.81) compared with 0 bouts/day. Daily MV-ILPA frequency showed a U-shaped pattern, with risk reductions plateauing at approximately 56 bouts/day (HR 0.54, 95% CI 0.39−0.76).

**Conclusions:** Among adults who do not do leisure-time exercise, accruing brief, intermittent bouts of moderate-to-vigorous intensity physical activity during day-to-day routines may be a promising strategy for prevention of type 2 diabetes.

## Background

Type 2 diabetes is a global public health concern, with its prevalence projected to increase from 5.9% in 2021 to 9.5% in 2050, affecting more than 1.27 billion people worldwide (1). Physical inactivity is a well-recognised modifiable risk factor, contributing at least 7.4% of global disability-adjusted life years (DALYs) attributable to type 2 diabetes in 2021 (1). Increasing regular physical activity, especially of moderate-to-vigorous intensity (MVPA), is therefore widely recommended as a first-line strategy for type 2 diabetes prevention (2,3).

Compared to other forms of physical activity, structured (leisure-time) exercise has long been regarded as the key behavioural strategy for preventing and managing type 2 diabetes (4,5). Despite its well-documented efficacy, evidence for long-term health effects is very limited, largely due to persistently low adherence in real-world settings (6). This reality supports the need to shift from an emphasis on exercise (which is planned and structured) towards a more holistic approach to physical activity promotion that better reflects real-world behaviour. In this context, incidental physical activity – defined as unstructured, unplanned activities of daily living performed as a by-product of an action with a different primary purpose (e.g., stair climbing, walking to the bus stop, gardening, carrying groceries/goods) (7) − may offer a practical alternative for the majority of the adult population (> 85%) (8,9) who do not engage in exercise regularly. This approach has strong potential to increase overall physical activity levels by leveraging behaviours that already make up the majority of daily movement. This is supported by a global survey showing that 88% of daily MVPA is accumulated through activities undertaken in the work, household and travel domains (10).

Most evidence to date on the beneficial health associations of PA has been derived from questionnaire-based data that typically captures only activity accrued through longer bouts (e.g., ≥ 10 minutes) and performed during leisure time (11). Recent advances in wearable technology and machine learning methods have enabled a more granular examination of physical activity “micropatterns”, such as activity accrued through brief and sporadic bursts of incidental activity occurring throughout the day (12,13). Specifically, a series of wearable-based studies using UK Biobank and National Health and Nutrition Examination Survey (NHANES) data have demonstrated steep beneficial health associations of vigorous intermittent lifestyle (i.e., incidental) physical activity (VILPA; bouts lasting up to 1 minute) (12,14–16) and its moderate-to-vigorous intensity equivalent (MV-ILPA; bouts lasting up to 3 minutes) (13). For example, accumulating 4.4 minutes/day or 3.0 bouts/day of VILPA was associated with 27% and 39% lower risk of all-cause mortality, respectively, compared with no VILPA (12). Similarly, all-cause mortality risk was 34% lower among individuals who accumulated MV-ILPA in bouts lasting 1 to < 3 minutes when compared with bouts of shorter than 1 minute (13). Micropatterns-accrued PA has not, however, been examined in relation to incident type 2 diabetes. Addressing this gap could help inform future clinical and public health guidelines by determining whether brief, incidental bursts of higher-intensity activity offer a practical and accessible strategy for preventing type 2 diabetes, particularly for individuals who are unable or unwilling to engage in leisure-time exercise.

The aim of this study was to examine the dose-response associations of daily duration and frequency of VILPA and MV-ILPA with incident type 2 diabetes in a large cohort of UK adults.

## Research Design and Methods

### Study Design and Participants

Data were drawn from the UK Biobank, a prospective population-based cohort of adults aged 40-69 years in the UK. Detailed descriptions of the study design and methods have been published elsewhere (17). Briefly, participants were recruited via postal invitations and attended baseline assessments between 2006 and 2010. Additionally, between June 2013 and December 2015, a subsample of 103,684 participants were mailed an Axivity AX3 accelerometer (Axivity Ltd, Newcastle Upon Tyne, UK) and instructed to wear it on their dominant wrist for seven consecutive days (24 hours/day) to measure physical activity (18). The UK Biobank received ethical approval from the National Research Ethics Service of the UK National Health Service (11/NW/0382), and all participants provided written informed consent.

For the present study, we included participants with valid accelerometry data (≥ 16 h/day of wear time) for at least three days, including at least one weekend day. As previously described (12,13,15), non-exerciser status was defined by excluding participants who reported engaging in leisure-time exercise or recreational walking more than once a week at baseline (see eTable 1 for the relevant questions). We further excluded participants who reported an inability to walk, had prevalent type 2 diabetes or were undergoing exogenous insulin therapy (ascertained from self-reports, primary care records, or hospital records) at or prior to accelerometry assessment, or had missing covariate data. To minimise reverse causation bias, participants who registered an event within the first two years of follow-up were excluded. The participant selection process is summarised in eFigure 1.

**Figure 1.**
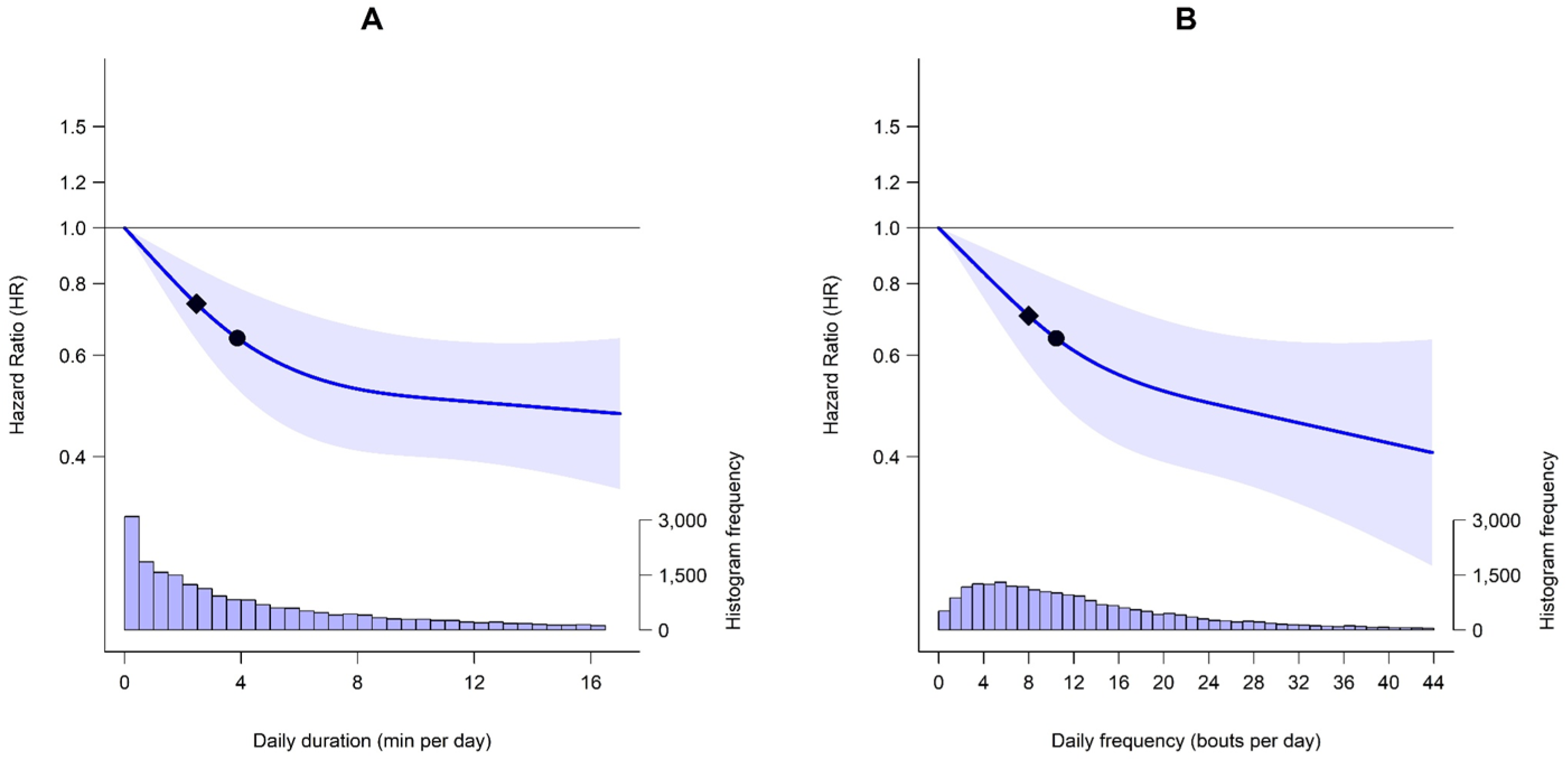
Multivariable-adjusted dose-response curves of daily vigorous intermittent lifestyle physical activity (VILPA) duration and frequency with incident type 2 diabetes (n = 22,706; events = 665). The line represents hazard ratios (HRs) and the shaded area represents their 95% confidence intervals associated with increasing daily (A) duration and (B) frequency of VILPA. The diamond refers to the minimal VILPA duration/frequency dose (as indicated by ED50 statistics) associated with 50% of optimal risk reduction, while the circle refers to the HR associated with the median duration/frequency of VILPA. The histogram on the right shows the sample distribution. The model was adjusted for sex, age, education levels, ethnicity, fruit and vegetable consumption, smoking status, alcohol consumption, sleep duration, discretionary screen time, medication use (cholesterol and blood pressure lowering), prevalent cancer, prevalent cardiovascular disease (CVD), parental history of cancer, CVD and type 2 diabetes (T2D), and physical activity energy expenditure volume of non-exposure intensity components (light-, moderate-and vigorous-intensity incidental physical activity [excluding VILPA]). The reference point was the minimum data point of VILPA duration (0 min/day) and frequency (0 bouts/day).

### Physical Activity Assessment and Processing

Prior to mailing, all AX3 accelerometers were initialised to collect data at a sampling frequency of 100 Hz with a dynamic range of ± 8 *g* (18). In this study, raw data files were calibrated, and non-wear time was identified using standard procedures (19,20). Physical activity was classified using a validated two-stage machine learning-based Random Forest activity classifier (84.6% accuracy across all four intensities) (21), as detailed in eMethod. Briefly, each 10-second window was first classified into one of four activity classes (sedentary, standing utilitarian movements, walking, or running/high-energy activities), which were then assigned to one of four intensity levels (sedentary, light, moderate, or vigorous).

We defined VILPA as brief bouts of vigorous intermittent lifestyle physical activity lasting up to 1 minute and MV-ILPA as the moderate-to-vigorous intermittent lifestyle physical activity lasting up to 3 minutes. The bout length selection was informed by: 1) laboratory data, where the average time to elicit VILPA was 76.7 [SD 6.8] seconds based on physiological responses (i.e., percentage of maximal oxygen uptake [VO_2max_] measured by indirect calorimetry and percentage of maximal heart rate) and perceived exertion (22); and 2) naturally occurring bout lengths observed in the UK Biobank sample, where 89.1% of incidental vigorous (15) and 87.2% of incidental moderate-to-vigorous activities (13) were accrued in bouts lasting up to 1 minute and 3 minutes, respectively.

### Type 2 Diabetes Ascertainment

The primary outcome for this study was incident type 2 diabetes, ascertained from multiple data sources (i.e., primary card records, hospital inpatient records and death registry) using Read codes version 2 and 3 for primary care records and the E11 code from the International Classification of Diseases 10th Revision (ICD-10) for hospital and death records (eTable 2). Participants were followed up through November 30, 2022, with deaths obtained via linkage with the National Health Service (NHS) Digital of England and Wales or the NHS Central Register and National Records of Scotland. Inpatient hospitalisation data were provided by the Hospital Episode Statistics for England, the Patient Episode Database for Wales, or the Scottish Morbidity Record for Scotland. Follow-up time was calculated as the time in years from accelerometry assessment to the first diagnosis of type 2 diabetes, death or censoring, whichever occurred first.

### Potential Confounders

Potential confounders were selected a priori based on previous analogue analyses using UK Biobank data (23,24). They included age, sex, ethnicity, education levels, smoking status, alcohol consumption, fruit and vegetable consumption, medication use (cholesterol and blood pressure lowering), previous diagnosis of cancer and CVD, parental history of cancer, CVD and type 2 diabetes, discretionary screen time, accelerometer-estimated sleep duration and physical activity energy expenditure (PAEE) volume for non-exposure intensity components (e.g., the analyses with VILPA duration and frequency as the exposure were adjusted for PAEE estimated from light-, moderate-and vigorous-intensity incidental activity from bouts lasting >1 minute). Complete covariate definitions are provided in eTable 3.

## Statistical Analysis

Participant characteristics were summarised by tertiles of daily MV-ILPA duration using means and standard deviations (SDs) or median and interquartile range (IQR) for continuous variables and counts and percentages for categorical variables. To minimise the influence of sparse data and outliers, the upper ranges of VILPA and MV-ILPA values were winsorised at the 97.5th percentile, and the lower range of MV-ILPA values was winsorised at the 2.5th percentile (12,14,15). We estimated the dose-response absolute risk between daily VILPA/MV-ILPA duration and frequency with incident type 2 diabetes (expressed as events per 10,000 person-years) using Poisson regression (25). Consistent with previous analogous analyses (13,14), we applied restricted cubic spline to demonstrate the dose-response curves with knots placed at the 10th, 50th and 90th percentiles of the exposures (26). We assessed time-to-event associations of daily VILPA/MV-ILPA duration and frequency with incident type 2 diabetes using Fine-Gray subdistribution hazard models to account for competing risks (27). Departure from linearity was assessed with a Wald test. Proportional hazard assumptions were tested using Schoenfeld residuals in the models, and no violations were observed (all p > 0.05). We calculated the minimum effective dose [ED50] (28) – defined as the daily VILPA/MV-ILPA duration and frequency associated with 50% of the optimal risk reduction – and reported the corresponding point estimates (hazard ratios [HRs] and 95% confidence intervals [CIs]) for median daily duration and frequency of VILPA/MV-ILPA. In all analyses, the minimum data point of the exposure was used as the reference (VILPA: 0 minute/day; MV-ILPA: 3.9 minutes/day).

We calculated E-values to assess the plausibility of bias from unmeasured confounding (29). To ensure the robustness of our findings, several sensitivity analyses were performed. We repeated main analyses with additional adjustment for cardiometabolic biomarkers that may be potential mediators: body mass index (BMI), waist circumference, low-density lipoprotein (LDL), high-density lipoprotein (HDL), triglycerides, and systolic and diastolic blood pressure. To further reduce the possibility of reverse causation bias, we excluded participants who were underweight (BMI < 18.5 kg/m^2^), reported poor self-rated health, or had a frailty index ≥ 3 (on a 0-5 scale) at baseline, and those who had prevalent cancer or CVD. We also tested the influence of analytical decisions by repeating the main analyses using the 25th percentile of the exposure distribution (VILPA: 1.3 minutes/day; MV-ILPA: 15.3 minutes/day) as the referent, and by applying an alternative spline specification with knots at the 10th, 33rd and 67th percentiles to account for potential data skewness. Finally, considering prior evidence of sex differences in the health associations of VILPA with CVD (15), we investigated potential effect modification by sex and conducted sex-stratified analyses of VILPA and MV-ILPA with incident type 2 diabetes.

All analyses were performed in R (version 4.5.1) using the rms (version 8.0-0) and survival (version 3.8.3) packages. This study was reported according to the Strengthening the Reporting of Observational Studies in Epidemiology (STROBE) guidelines (eTable 4).

## Data Availability Statement

UK Biobank data were obtained under application number 25813. UK Biobank data are available to researchers with an approved request (https://www.ukbiobank.ac.uk/register-apply/).

## Results

### Participant Characteristics

The final analytical sample comprised 22,706 non-exercising adults (mean age 62.3 [SD 7.6] years; 56.4% female), with 665 incident type 2 diabetes occurring over an average follow-up period of 7.9 years (SD 1.0). The sample was predominantly White (99.1%), with 37% having a college or university degree, 55.7% having never smoked, and 59.3% reporting alcohol consumption within current UK guidelines (≤ 14 units per week). Daily median (IQR) duration and frequency were 3.9 (1.5–8.9) minutes/day and 10.4 (5.4−17.9) bouts/day for VILPA, and 25.3 (15.3–40.8) minutes/day and 39.0 (25.6−57.4) bouts/day for MV-ILPA, respectively.

Participant characteristics by tertiles of daily MV-ILPA duration are presented in Table 1.

**Table 1.** Participant characteristics by tertiles of daily moderate-to-vigorous intermittent lifestyle physical activity (MV-ILPA) duration.

|  | Tertiles of daily MV-ILPA duration |  |  | Full sample |
| --- | --- | --- | --- | --- |
|  | 3.8-18.4 min/day | 18.4-34.4 min/day | > 34.4 min/day |  |
| **Sample size** | 7561 | 7562 | 7583 | 22,706 |
| **Follow up in years** | 7.8 (1.2) | 7.9 (1.0) | 8.0 (0.9) | 7.9 (1.0) |
| **Age in years** | 63.9 (7.4) | 62.1 (7.6) | 60.8 (7.6) | 62.3 (7.6) |
| **Male, n (%)** | 3,213 (42.5) | 3,298 (43.6) | 3,397 (44.8) | 9,908 (43.6) |
| **Ethnicity: non-White, n (%)** | 67 (0.9) | 66 (0.9) | 57 (0.8) | 190 (0.9) |
| **Education levels, n (%)** |  |  |  |  |
| College or university degree | 2,890 (38.2) | 2,950 (39.0) | 2,708 (35.7) | 8,548 (37.6) |
| A/AS level | 1,013 (13.4) | 905 (12.0) | 948 (12.5) | 2,866 (12.6) |
| General certificate of secondary education | 1,604 (21.2) | 1,693 (22.4) | 1,754 (23.1) | 5,051 (22.2) |
| Certificate of secondary education | 320 (4.2) | 331 (4.4) | 499 (6.6) | 1,150 (5.1) |
| National Vocational Qualifications, Higher National Diploma, or Higher National Certificate | 464 (6.1) | 480 (6.3) | 482 (6.4) | 1,426 (6.3) |
| Other | 1,270 (16.8) | 1,203 (15.9) | 1,192 (15.7) | 3,665 (16.1) |
| **Smoking status, n (%)** |  |  |  |  |
| Current | 816 (10.8) | 644 (8.5) | 604 (8.0) | 2,064 (9.1) |
| Never | 4,038 (53.4) | 4,264 (56.4) | 4,349 (57.4) | 12,651 (55.7) |
| Previous | 2,707 (35.8) | 2,654 (35.1) | 2,630 (34.7) | 7,991 (35.2) |
| **Alcohol consumption\*, n (%)** |  |  |  |  |
| Never | 304 (4.0) | 279 (3.7) | 268 (3.5) | 851 (3.7) |
| Ex-drinker | 258 (3.4) | 252 (3.3) | 234 (3.1) | 744 (3.3) |
| Within guidelines | 4,497 (59.5) | 4,503 (59.5) | 4,475 (59.0) | 13,475 (59.3) |
| Above guidelines | 2,502 (33.1) | 2,528 (33.4) | 2,606 (34.4) | 7,636 (33.6) |
| **Fruit and vegetable consumption (servings/day)** | 7.4 (4.3) | 7.3 (4.1) | 7.5 (4.2) | 7.4 (4.2) |
| **Medication, n (%)** |  |  |  |  |
| Cholesterol lowering | 1,430 (18.9) | 1,134 (15.0) | 866 (11.4) | 3,430 (15.1) |
| Blood pressure lowering | 1,824 (24.1) | 1,403 (18.6) | 1,097 (14.5) | 4,324 (19.0) |
| **Prevalent cancer, n (%)** | 689 (9.1) | 604 (8.0) | 487 (6.4) | 1,780 (7.8) |
| **Prevalent cardiovascular disease, n (%)** | 1,121 (14.8) | 881 (11.7) | 683 (9.0) | 2,685 (11.8) |
| **Parental history of cancer, n (%)** | 2,671 (35.3) | 2,594 (34.3) | 2,348 (31.0) | 7,613 (33.5) |
| **Parental history of cardiovascular disease, n (%)** | 4,515 (59.7) | 4,419 (58.4) | 4,181 (55.1) | 13,115 (57.8) |
| **Parental history of type 2 diabetes, n (%)** | 1,277 (16.9) | 1,271 (16.8) | 1,288 (17.0) | 3,836 (16.9) |
| **Body mass index (kg/m<sup>2</sup>)** | 28.4 (5.2) | 27.3 (4.7) | 26.5 (4.4) | 27.4 (4.8) |
| **Waist circumference (cm)** | 92.9 (13.9) | 90.1 (13.2) | 88.0 (12.6) | 90.3 (13.4) |
| **Biomarkers** |  |  |  |  |
| High-density lipoprotein (mmol/L) | 1.4 (0.4) | 1.4 (0.4) | 1.5 (0.4) | 1.4 (0.4) |
| Low-density lipoprotein (mmol/L) | 3.6 (0.9) | 3.6 (0.8) | 3.6 (0.8) | 3.6 (0.9) |
| Triglycerides (mmol/L) | 1.8 (1.0) | 1.8 (1.0) | 1.7 (1.0) | 1.8 (1.0) |
| Systolic blood pressure (mmHg) | 141.2 (19.6) | 138.8 (18.8) | 138.1 (18.8) | 139.4 (19.1) |
| Diastolic blood pressure (mmHg) | 83.2 (10.7) | 82.3 (10.6) | 81.9 (10.5) | 82.5 (10.6) |
| **Self-rated health: Poor, n (%)** | 434 (5.7) | 286 (3.8) | 218 (2.9) | 938 (4.1) |
| **High frailty, n (%)†** | 441 (6.2) | 242 (3.4) | 210 (2.9) | 893 (4.2) |
| **Discretionary screen time (hours/day)** | 4.0 [3.0, 5.5] | 4.0 [2.5, 5.0] | 3.5 [2.5, 5.0] | 4.0 [2.5, 5.0] |
| **Sleep duration (min/day), median [IQR]** | 441.0 [396.8, 480.9] | 439.1 [397.6, 475.9] | 437.3 [397.7, 473.3] | 439.1 [397.4, 476.5] |
| **Sedentary behaviour (min/day), median [IQR]** | 696.6 [647.3, 749.6] | 656.0 [606.8, 706.1] | 598.4 [544.9, 649.9] | 651.5 [593.4, 710.2] |
| **Physical activity (min/day)** |  |  |  |  |
| Light intensity, median [IQR] | 68.7 [49.7, 99.3] | 90.9 [62.8, 132.8] | 128.2 [82.9, 179.4] | 91.4 [61.1, 140.1] |
| Moderate intensity, median [IQR] | 10.3 [6.7, 13.7] | 23.0 [19.1, 27.7] | 46.4 [36.7, 63.6] | 23.3 [13.5, 38.2] |
| Vigorous intensity, median [IQR] | 1.4 [0.5, 3.5] | 4.2 [1.9, 9.0] | 8.5 [3.6, 18.1] | 3.9 [1.3, 9.6] |
| VILPA, median [IQR] | 1.4 [0.5, 3.2] | 4.0 [1.8, 8.0] | 7.8 [3.5, 15.4] | 3.9 [1.5, 8.9] |
| MV-ILPA, median [IQR] | 11.8 [8.0, 15.2] | 25.3 [21.7, 29.5] | 49.7 [40.8, 66.6] | 25.3 [15.3, 40.8] |
| **Physical activity (bouts/day)** |  |  |  |  |
| VILPA, median [IQR] | 4.9 [2.6, 8.1] | 10.9 [7.0, 15.9] | 19.2 [12.5, 28.3] | 10.4 [5.4, 17.9] |
| MV-ILPA, median [IQR] | 20.9 [14.9, 26.7] | 39.3 [33.9, 45.6] | 67.0 [55.9, 84.6] | 39.0 [25.6, 57.4] |
| **Physical activity energy expenditure (kJ/kg/day)** |  |  |  |  |
| Light intensity, median [IQR] | 6.5 [4.8, 8.9] | 9.1 [6.7, 12.3] | 13.4 [9.4, 17.4] | 9.1 [6.4, 13.3] |
| Moderate intensity, median [IQR] | 1.8 [1.2, 2.4] | 4.1 [3.4, 5.0] | 8.5 [6.6, 11.6] | 4.2 [2.4, 7.0] |
| Vigorous intensity, median [IQR] | 0.4 [0.2, 0.8] | 1.1 [0.6, 1.9] | 2.1 [1.1, 3.6] | 1.0 [0.4, 2.1] |
| **Type 2 diabetes incidence, n (%)** | 309 (4.1) | 202 (2.7) | 154 (2.0) | 665 (2.9) |

Abbreviations: IQR, interquartile range; VILPA, vigorous intermittent lifestyle physical activity; MV-ILPA, moderate-to-vigorous intermittent
lifestyle physical activity. \*Defined based on UK guidelines of $\leq 14$ units of alcohol per week (1 unit = 8g of pure ethanol). †Frailty index of $\geq 3$ (on a scale of 0 to 5).
Data are presented as mean (standard deviation), unless otherwise indicated.

### Dose-response Association of VILPA and Type 2 Diabetes Incidence

The multivariable-adjusted models of absolute risk (eFigure 2A) and relative HR (Figure 1A) showed that daily VILPA duration was associated with lower type 2 diabetes risk in an L-shaped manner. The median daily VILPA duration (3.9 minutes/day) was associated with an absolute risk of 15.2 (95% CI 11.7−19.7) per 10,000 person-years (eFigure 2A). Compared to the minimum referent data point (0 minutes/day), we observed a non-linear dose-response association (p_non-linear_ = 0.007) between daily VILPA duration and incident type 2 diabetes, with the curve plateauing at approximately 8.0 minutes/day (HR 0.52; 95% CI 0.41–0.67) (Figure 1A). The minimum effective dose (ED50) of VILPA duration was 2.5 minutes/day, corresponding to a HR of 0.73 (95% CI 0.64–0.85). The median daily VILPA duration was associated with a HR of 0.64 (95% CI 0.52–0.79).

**Figure 2.**
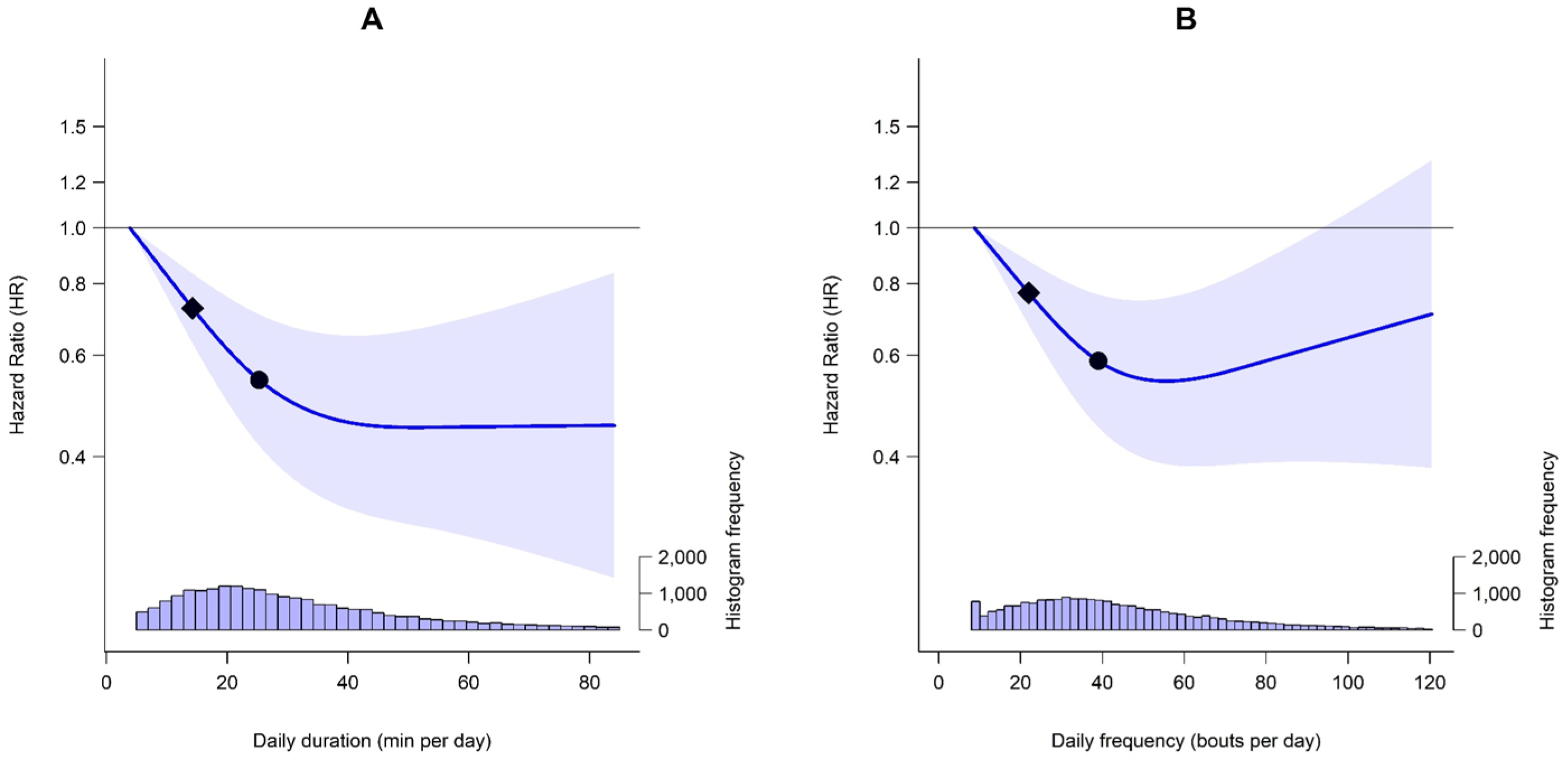
Multivariable-adjusted dose-response curves of daily moderate-to-vigorous intermittent lifestyle physical activity (MV-ILPA) duration and frequency with incident type 2 diabetes (n = 22,795; events = 751). The line represents hazard ratios (HRs) and the shaded area represents their 95% confidence intervals associated with increasing daily (A) duration and (B) frequency of MV-ILPA. The diamond refers to the minimal MV-ILPA duration/frequency dose (as indicated by ED50 statistics) associated with 50% of optimal risk reduction, while the circle refers to the HR associated with the median duration/frequency of MV-ILPA. The histogram on the right shows the sample distribution. The model was adjusted for sex, age, education levels, ethnicity, fruit and vegetable consumption, smoking status, alcohol consumption, sleep duration, discretionary screen time, medication use (cholesterol and blood pressure lowering), prevalent cancer, prevalent cardiovascular disease (CVD), parental history of cancer, CVD and type 2 diabetes (T2D), and physical activity energy expenditure volume of non-exposure intensity components (light-, moderate-and vigorous-intensity incidental physical activity [excluding MV-ILPA]). The reference point was the minimum data point of MV-ILPA duration (3.9 min/day) and frequency (8.7 bouts/day).

Daily VILPA frequency showed a more linear dose-response association with incident type 2 diabetes compared with the association of daily duration. The median daily VILPA frequency (10.4 bouts/day) was associated with an absolute risk of 15.9 (95% CI 12.4−20.5) per 10,000 person-years (eFigure 2B). Compared to the minimum referent data point (0 bouts/day), there was a near-linear dose-response association (p_non-linear_ = 0.060) with lower incident type 2 diabetes (Figure 1B). The minimum effective dose (ED50) of daily VILPA frequency was 8.0 bouts/day (HR 0.70, 95% CI 0.58−0.85), and the median daily frequency was associated with a HR of 0.64 (05% CI 0.51−0.81).

### Dose-response Association of MV-ILPA and Type 2 Diabetes Incidence

Daily MV-ILPA duration showed a similar dose-response pattern to VILPA duration, but with a steeper slope. The median daily MV-ILPA duration (25.3 minutes/day) was associated with an absolute risk of 15.2 (95% CI 11.8−19.6) per 10,000 person-years (eFigure 3A). Compared to the minimal referent data point (3.9 minutes/day), daily MV-IPA duration was non-linearly associated with incident type 2 diabetes (p_non-linear_ = 0.003), with the curve plateauing at approximately 40 minutes/day (HR 0.46, 95% CI 0.32–0.65) (Figure 2A). The minimum effective dose (ED50) of MV-ILPA duration was 14.2 minutes/day, corresponding to a HR of 0.73 (95% CI 0.63–0.83). The median daily MV-ILPA duration was associated with a HR of 0.54 (95% CI 0.42−0.71).

For daily MV-ILPA frequency, both absolute (eFigure 3B) and relative risk (Figure 2B) analyses demonstrated a U-shaped non-linear dose-response association with incident type 2 diabetes. The median daily MV-ILPA frequency (39.0 bouts/day) was associated with an absolute risk of 15.2 (95% CI 11.8−19.6) per 10,000 person-years (eFigure 3B). Compared to the minimum referent data point (8.7 bouts/day), the non-linear dose-response curve for MV-ILPA frequency (p_non-linear_ < 0.001) began to plateau at approximately 56 bouts/day – the data point corresponding to the lowest estimate (HR 0.54, 95% CI 0.39−0.76) – followed by a gradual upward slope at higher frequencies (Figure 2B). The minimum effective dose (ED50) of MV-ILPA frequency was 21.9 bouts/day (HR 0.77, 95% CI 0.68-0.86), and the median daily frequency was associated with a HR of 0.59 (95% CI 0.45−0.77).

## Sensitivity Analyses

The E-values ranged from 1.78 (lower 95% CI 1.48) to 2.05 (1.63) for VILPA and from 1.69 (1.42) to 2.42 (1.86) for MV-ILPA, suggesting that a moderate degree of unmeasured confounding would be required to nullify the observed associations with incident type 2 diabetes (eTable 5). Additional adjustment for BMI, waist circumference and other cardiometabolic biomarkers showed similar results for VILPA (eFigures 4 and 5) but attenuated dose-response associations for MV-ILPA (eFigures 6 and 7) compared with the main analyses. Excluding participants who were underweight or reported poor self-rated health or had a high frailty index resulted in clearer and slightly steeper dose-response associations for both VILPA (eFigures 8 and 9) and MV-ILPA (eFigures 10 and 11) with type 2 diabetes incidence. Similarly, excluding participants with prevalent cancer or CVD produced clearer and steeper dose-response curves for both VILPA (eFigure 12) and MV-ILPA (eFigure 13), with VILPA frequency showing a more linear pattern. When setting the referent to the 25th percentile of exposure, the association pattern remained consistent for both VILPA (eFigure 14) and MV-ILPA (eFigure 15). Using alternative knot placements generated similar dose-response curves, but with a stronger magnitude of association for both exposures compared with the main analyses (eFigures 16 and 17). Finally, there was only weak evidence of effect modification by sex for VILPA (p_interaction_ for duration and frequency = 0.334 and 0.364, respectively; eFigure 18) and MV-ILPA (p_interaction_ for duration and frequency = 0.823 and 0.682, respectively; eFigure 19).

## Conclusions

To our knowledge, this is the first study to investigate the associations of physical activity micropatterns with incident type 2 diabetes. We found that both daily duration of VILPA and MV-ILPA were inversely associated with incident type 2 diabetes in a L-shaped manner, with the sample median durations (VILPA: 3.9 minutes/day; MV-ILPA: 25.3 minutes/day) corresponding to 36% and 46% lower risk of incident type 2 diabetes, respectively. We also found a near-linear inverse association between daily frequency of VILPA and type 2 diabetes incidence, indicating progressively lower risk with increasing numbers of bouts taken daily. In contrast, daily frequency of MV-ILPA was inversely associated with incident type 2 diabetes in a U-shaped manner, with the strongest risk reduction (46%) observed at approximately 55.7 bouts/day. Our sensitivity analyses showed that these associations were broadly consistent across a range of measures against reverse causation (e.g., excluding prevalent cancer and CVD; setting the 25th percentile as referent data point) and other analytical assumptions (e.g., using different reference points or knot placements). Collectively, these findings suggest that brief bursts of higher-intensity incidental physical activity integrated into daily routines may be a time-efficient and potentially more behaviourally sustainable promising approach for prevention of type 2 diabetes.

Current type 2 diabetes prevention and management guidelines tend to focus on structured exercise as the primary behavioural intervention (4), despite its persistently low uptake and adherence in real-world settings (6). Our analyses showed that regular brief bouts of moderate-to-vigorous activity embedded within daily routines may offer comparable benefits for individuals who do not formally engage in structured exercise. Specifically, accumulating VILPA equivalent to the median of 3.9 minutes/day (i.e., 36.4% of the WHO’s weekly recommended duration of vigorous physical activity) (2) or 10.4 bouts/day was associated with a 36% lower risk of developing type 2 diabetes compared to those who completed no VILPA. Furthermore, the near-linear pattern observed with daily VILPA frequency may indicate that integrating brief, frequent VILPA bouts into daily routines could offer more consistent benefits than accumulating longer daily durations alone in the context of type 2 diabetes prevention. The observed lower type 2 diabetes risk associated with greater micropatterns-accrued PA may be partly explained by analogous physiological adaptations as those elicited by structured exercise, including maintenance and improvement of cardiorespiratory fitness, insulin sensitivity and glycaemic control (30,31).

Consistent with the dose-response pattern observed for VILPA, daily MV-ILPA duration showed a similar non-linear trend, with the median duration of 25.3 minutes/day associated with a 46% lower risk of type 2 diabetes. In contrast to VILPA frequency, we found a U-shaped association between daily MV-ILPA frequency and incident type 2 diabetes, suggesting that excessively high number of MV-ILPA bouts (beyond ∼56 bouts/day) may not be associated with further risk reduction. The observed beneficial associations with MV-ILPA could possibly be due to moderate-intensity activity eliciting physiological adaptations (e.g., improved metabolic flexibility) that complement those induced by vigorous-intensity activity (e.g., activation of glycolytic pathways) (32,33), which together may strengthen insulin sensitivity and overall cardiometabolic function. Notably, the minimum effective dose (i.e., 50% of the total effect size [ED50]) of MV-ILPA duration (14.2 minutes/day) corresponded to the same type 2 diabetes risk reduction (27%) as those of VILPA duration (2.5 minutes/day), suggesting that comparable risk reductions may be achieved through longer daily durations of incidental activity at moderate-to-vigorous intensity. This finding aligns with previous health equivalence analyses showing that each minute of incidental activity accrued at vigorous intensity may offer CVD risk reductions equivalent to 2.8-3.4 minutes of moderate-intensity incidental activity (21). This demonstrates the flexibility in how incidental activity can be accumulated across different intensities with similar protective effects, providing a range of feasible and potentially behaviourally sustainable options for type 2 diabetes prevention.

Our findings are consistent with previous literature showing substantial risk reductions (∼16-45% lower risk) for VILPA and MV-ILPA across multiple health outcomes, including all-cause mortality (12,16), cancer mortality (12) and incidence (14) and major adverse cardiovascular events (13,15). Collectively, this evidence highlights the potential of incidental physical activity to offer a practical alternative for adults who do not exercise regularly, with previous qualitative work indicating that VILPA aligns well with individuals’ daily routines and priorities (34). Further, recent pilot randomised controlled trials demonstrated the feasibility and acceptability of a micropattern-focused intervention among adults transitioning into retirement (35) and women from socioeconomically diverse backgrounds (36), with improvements observed in daily incidental physical activity levels. While larger-scale trials are needed, these preliminary findings suggest the integration of brief bursts of vigorous-and/or moderate-intensity incidental physical activity into everyday life is a promising alternative strategy in type 2 diabetes prevention. Importantly, as our findings are based on *daily averages* of VILPA and MV-ILPA, the observed risk reductions likely reflect consistent day-to-day accumulation patterns, reinforcing the importance of habitual physical activity in preventing noncommunicable disease (2).

The strengths of our study include the use of high-resolution accelerometer data to capture physical activity micropatterns and the unique focus on non-exercising adults, which enabled investigation of the health associations of short bouts of intermittent physical activity embedded into daily living. The use of a validated two-stage machine learning-based activity classifier also improved the precision of incidental activity type and intensity classification (21), although underestimation of VILPA/MV-ILPA remains possible due to the inherent limitations of wrist-worn accelerometers in capturing certain activities (e.g., walking uphill, or while carrying heavy loads). Despite the long follow-up period and extensive sensitivity analyses, the possibility of reverse causation bias cannot be fully ruled out. There was a median lag of 5.5 years between the baseline questionnaire data collection (including self-report leisure-time exercise and other covariates) and accelerometry measurements; however, previous studies have shown that non-exerciser status (82-88% retained this status based on re-examination data) (12) and most covariates (37) were generally stable over time. While the UK Biobank had a very low response rate (5.5%) and may be susceptible to a ‘healthy volunteer’ selection bias (i.e., participants were generally healthier and more socioeconomically advantaged than the broader UK population), the heterogeneity of exposures still allow for valid inferences about exposure-health associations that may be generalisable to other populations, as shown by previous empirical work (38). Finally, given the observational design, our findings cannot provide definitive evidence of the causality of the observed associations.

In summary, we found that micropatterns-accrued incidental PA of higher intensities was consistently associated with lower type 2 diabetes incidence in a dose-response manner. These findings support the promotion of short, regular bouts of incidental physical activity at moderate-and/or vigorous-intensity as part of daily routines, offering a promising strategy for type 2 diabetes prevention.

## Acknowledgements

This research has been conducted using the UK Biobank Resource under Application Number 25813. The authors would like to thank all the participants and professionals contributing to the UK Biobank.

## Funding and Assistance

This study is funded by an Australian National Health and Medical Research Council (NHMRC) Investigator Grant (APP1194510) and the National Heart Foundation of Australia (APP107158). The funder had no specific role in any of the following study aspects: the design and conduct of the study; collection, management, analysis, and interpretation of the data; preparation, review, or approval of the manuscript; and decision to submit the manuscript for publication.

## Author Contributions and Guarantor Statement

K.H.C., M.N.A., R.K.B., N.A.K. and E.S. contributed to the conceptualisation of the analysis plan. M.N.A. and R.K.B. contributed to the derivation of data variables. K.H.C. conducted the formal analyses. All authors contributed to the interpretation of the results. K.H.C. wrote the first and subsequent drafts of the manuscript under the guidance of E.S. and M.N.A., and all other authors edited, reviewed, and approved the final version of the manuscript. K.H.C. and E.S. are the guarantors of this work and, as such, had full access to all the data in the study and take responsibility for the integrity of the data and the accuracy of the data analysis.

