## Supplementary Material for "Dose-response associations of intermittent lifestyle physical activity micropatterns and incident type 2 diabetes"

### TABLE OF CONTENTS


| Page | Item |
|---|---|
| 3 | **eTable 1:** List of questions to access participation in leisure time physical activity |
| 5 | **eTable 2:** Definitions of type 2 diabetes |
| 6 | **eTable 3:** Definitions of covariates |
| 8 | **eTable 4:** STROBE Statement |
| 11 | **eTable 5:** E-values for minimum effective dose and median VILPA and MV-ILPA values for incident type 2 diabetes |
| 12 | **eFigure 1:** Participant selection process |
| 13 | **eFigure 2:** Adjusted absolute risk-based dose response curves of daily VILPA duration and frequency with incident type 2 diabetes (n = 22,706; events = 665) |
| 14 | **eFigure 3:** Adjusted absolute risk-based dose response curves of daily MV-ILPA duration and frequency with incident type 2 diabetes (n = 22,706; events = 665) |
| 15 | **eFigure 4:** Adjusted dose response curves of daily VILPA duration and frequency with incident type 2 diabetes, with additional adjustment for BMI and other cardiometabolic biomarkers (n = 18,313; events = 526) |
| 16 | **eFigure 5:** Adjusted dose response curves of daily VILPA duration and frequency with incident type 2 diabetes, with additional adjustment for waist circumference and other cardiometabolic biomarkers (n = 18,554; events = 533) |
| 17 | **eFigure 6:** Adjusted dose response curves of daily MV-ILPA duration and frequency with incident type 2 diabetes, with additional adjustment for BMI and other cardiometabolic biomarkers (n = 18,313; events = 526) |
| 18 | **eFigure 7:** Adjusted dose response curves of daily MV-ILPA duration and frequency and incident type 2 diabetes, with additional adjustment for waist circumference other cardiometabolic biomarkers (n = 18,554; events = 533) |
| 19 | **eFigure 8:** Adjusted dose response curves of daily VILPA duration and frequency and incident type 2 diabetes, excluding participants who were underweight (BMI < 18.5 kg/m2) or reported poor health (n = 21,346; events = 584) |
| 20 | **eFigure 9:** Adjusted dose response curves of daily VILPA duration and frequency with incident type 2 diabetes, excluding participants with a frailty index score of 3 or higher (on a 0-5 scale) (n = 20,435; events = 570) |

| | |
|---|---|
| **21** | **eFigure 10:** Adjusted dose response curves of daily MV-ILPA duration and frequency and incident type 2 diabetes, excluding participants who were underweight (BMI < 18.5 kg/m2) or reported poor health (n = 21,346; events = 584) |
| **22** | **eFigure 11:** Adjusted dose response curves of daily MV-ILPA duration and frequency with incident type 2 diabetes, excluding participants with a frailty index score of 3 or higher (on a 0-5 scale) (n = 20,435; events = 570) |
| **23** | **eFigure 12:** Adjusted dose response curves of daily VILPA duration and frequency with incident type 2 diabetes, excluding participants with prevalent cancer or CVD (n = 18,380; events = 476) |
| **24** | **eFigure 13:** Adjusted dose response curves of daily MV-ILPA duration and frequency with incident type 2 diabetes, excluding participants with prevalent cancer or CVD (n = 18,380; events = 476) |
| **25** | **eFigure 14:** Adjusted dose response curves of daily VILPA duration and frequency and incident type 2 diabetes using the 25th percentile of VILPA duration as the reference point (n = 22,706; events = 665) |
| **26** | **eFigure 15:** Adjusted dose response curves of daily MV-ILPA duration and frequency and incident type 2 diabetes using the 25th percentile of MV-ILPA duration as the reference point (n = 22,706; events = 665) |
| **27** | **eFigure 16:** Adjusted dose response curves of daily VILPA duration and frequency and incident type 2 diabetes using alternative knots placement (10th, 33rd and 67th percentiles) (n = 22,706; events = 665) |
| **28** | **eFigure 17:** Adjusted dose response curves of daily MV-ILPA duration and frequency and incident type 2 diabetes using alternative knots placement (10th, 33rd and 67th percentiles) (n = 22,706; events = 665) |
| **29** | **eFigure 18:** Sex-specific adjusted dose response curves of daily VILPA duration and frequency and incident type 2 diabetes (male: n = 9,908; events = 400; female, n = 12,798; events = 265) |
| **30** | **eFigure 19:** Sex-specific adjusted dose response curves of daily MV-ILPA duration and frequency and incident type 2 diabetes (male: n = 9,908; events = 400; female, n = 12,798; events = 265) |
| **31** | **eMethod:** Physical activity classification |
| **35** | **References** |

**eTable 1:** List of questions to access participation in leisure time physical activity

| No. | Questions | Response options |
|---|---|---|
| 1 | In the last 4 weeks, did you spend any time doing the following? (You can select more than one answer)<br><br>Strenuous sports include sports that make you sweat or breathe hard. Heavy DIY includes chopping wood, home or car maintenance, lifting heavy objects or using heavy tools *(UK Biobank field ID: 6164)* | • Walking for pleasure (not as a means of transport)<br>• Other exercises (eg: swimming, cycling, keep fit, bowling)<br>• Strenuous sports<br>• Light DIY (eg: pruning, watering the lawn)<br>• Heavy DIY (eg: weeding, lawn mowing, carpentry, digging)<br>• None of the above<br>• Prefer not to answer |
| 2 | Each time you did strenuous sports, about how long did you spend doing it? *(UK Biobank field ID: 1001)* | • Less than 15 minutes<br>• Between 15 and 30 minutes<br>• Between 30 minutes and 1 hour<br>• Between 1 hour and 1.5 hours<br>• Between 1.5 hours and 2 hours<br>• Between 2 and 3 hours<br>• Over 3 hours<br>• Do not know<br>• prefer not to answer |
| 3 | How many times in the last 4 weeks did you do strenuous sports? *(UK Biobank field ID: 991)* | • Once in the last 4 weeks<br>• 2-3 times in the last 4 weeks<br>• Once a week<br>• 2-3 times a week<br>• 4-5 times a week<br>• Every day<br>• Do not know<br>• Prefer not to answer |
| 4 | Each time you did other exercises such as swimming, cycling, keep fit, about how long did you spend doing it? *(UK Biobank field ID: 3647)* | • Less than 15 minutes<br>• Between 15 and 30 minutes<br>• Between 30 minutes and 1 hour<br>• Between 1 hour and 1.5 hours<br>• Between 1.5 hours and 2 hours<br>• Between 2 and 3 hours |

| | | |
|---|---|---|
| 5 | How many times in the last 4 weeks did you do other exercises such as swimming, cycling, keep fit? *(UK Biobank field ID: 3637)* | • Once in the last 4 weeks<br>• 2-3 times in the last 4 weeks<br>• Once a week<br>• 2-3 times a week<br>• 4-5 times a week<br>• Every day<br>• Do not know<br>• Prefer not to answer |
| 6 | Each time you went walking for pleasure, about how long did you spend doing it? *(UK Biobank field ID: 981)* | • Less than 15 minutes<br>• Between 15 and 30 minutes<br>• Between 30 minutes and 1 hour<br>• Between 1 hour and 1.5 hours<br>• Between 1.5 hours and 2 hourS<br>• Between 2 and 3 hours<br>• Over 3 hours<br>• Do not know<br>• prefer not to answer |
| 7 | How many times in the last 4 weeks did you go walking for pleasure? *(UK Biobank field ID: 971)* | • Once in the last 4 weeks<br>• 2-3 times in the last 4 weeks<br>• Once a week<br>• 2-3 times a week<br>• 4-5 times a week<br>• Every day<br>• Do not know<br>• Prefer not to answer |

Note. Participation in exercise and sport was measured through a close-ended questionnaire that asked participants to report the frequency and duration they engaged in exercise or sport. Participants who did not indicate engaging in exercise or sport (Question 1) were not given the option to answer Questions 2-5. Only participants who indicated no engagement in exercise or sport and walked for pleasure one time or less a week were included in the present analyses.

**eTable 2:** Definitions of type 2 diabetes

| **Variable** | **Definitions** |
|---|---|
| Type 2 diabetes | ICD-10 code using death register and inpatient hospitalisation: E11.<br><br>Read Codes using general practitioner records:<br>Version 2: C1041, C1096, C109A, C109B, C109C, C109E, C109F, C109G, C109H, C10F6, C10FA, C10FB, C10FC, C10FE, C10FF, C10FG, C10FH, C10FL, C10FM, C10FQ, C10FR.<br><br>Version 3: C1011, C102, C1021, C1031, C1041, C1051, C1061, C1071, C1074, C1090, C1091, C1092, C1093, C1094, C1095, C1096, C1097, C10y1, C10z1, X40J5, X40J6, X40JJ, Xaagf, XaCJ2, XaELQ, XaEnp, XaEnq, XaF05, XaFmA, XaFn7, XaFn8, XaFn9, XaFWI, XaIrf, XaIzQ, XaIzR, XaJQp, XaKyX, XE10F, XSETH. |

**eTable 3:** Definitions of covariates

| Variable | Definition | UK Biobank field ID (if applicable) |
|---|---|---|
| **Main analyses** | | |
| Age | Continuous | 34, 52, accelerometer date-timestamp |
| Sex | Female/Male | 31 |
| Ethnicity | White/Others | |
| Education levels | College/University; A/AS level; O levels; CSE; NVQ/HND/HNC; other | 6138 |
| Smoking status | Never, past, current | 20116 |
| Alcohol consumption | Never, ex-drinker, within guidelines, above guidelines | 20117, 1558 |
| Sleep duration | Hours spent sleeping | Derived from accelerometer data |
| Fruit and vegetable consumption | Continuous; number of servings/day | 1309, 1319, 1289, 1299 |
| Discretionary screen time | Calculated as the sum of TV viewing time plus (non-occupational) leisure time computer use (hours/day).<br><br>Participants were asked: 'In a typical day, how many hours do you spend watching TV?' They were also asked about time spent using a computer: 'In a typical day, how many hours do you spend using the computer? (Do not include using a computer at work)'. | 1070, 1080 |
| Prevalent cancer | Identified by self-report and cancer registry | 20001, 100092 |
| Prevalent CVD | Identified by self-report and hospitalisation | 20002, 41270 |
| Parental history of CVD, cancer and type 2 diabetes | Self-reported mother or father diagnosed with diseases | 20107, 20110 |
| Use of cholesterol medication | Yes/No | 6177, 6153 |

| | | |
|---|---|---|
| Use of blood pressure lowering medication | Yes/No | 6177, 6153 |
| PAEE volume (light-intensity physical activity [1.5 to < 3 METs)† | Continuous; kJ/kg/day | Derived from accelerometer data |
| PAEE volume (moderate-intensity physical activity [≥ 3 to < 6 METs)† | Continuous; kJ/kg/day | Derived from accelerometer data |
| PAEE volume (vigorous-intensity physical activity [≥ 6 METs)§ | Continuous; kJ/kg/day (excluding VILPA) | Derived from accelerometer data |
| PAEE volume (moderate- to vigorous-intensity physical activity) | Continuous; kJ/kg/day (excluding VILPA) | Derived from accelerometer data |
| **Sensitivity analyses** | | |
| Body mass index | Continuous; $kg/m^2$ | 23104 |
| Waist Circumference | Continuous; cm | 48 |
| Low density lipoprotein | Continuous; mmol/mol | 30780 |
| High density lipoprotein | Continuous; mmol/mol | 30760 |
| Triglycerides | Continuous; mmol/L | 30870 |
| Systolic blood pressure | Continuous; mmHg | 4080 |
| Diastolic blood pressure | Continuous; mmHg | 4079 |
| High frailty | Yes/No; high frailty indicates a score of ≥3 on a 0 to 5 scale | 2306, 120107, 2624, 1011, 3637, 991, 971, 924, 46, 47 |
| Self-rated health | Poor | 2178 |

Abbreviations: CVD, cardiovascular disease; PAEE, physical activity energy expenditure.

†Estimated using an established method by White et al[1], which has been validated against doubly labelled water to estimate instantaneous PAEE from the accelerometer data.

**eTable 4:** STROBE statement

| | Item No | Recommendation | Page No |
|---|---|---|---|
| **Title and abstract** | 1 | (*a*) Indicate the study's design with a commonly used term in the title or the abstract | 3 |
| | | (*b*) Provide in the abstract an informative and balanced summary of what was done and what was found | 3-4 |
| **Introduction** | | | |
| Background/rationale | 2 | Explain the scientific background and rationale for the investigation being reported | 6-7 |
| Objectives | 3 | State specific objectives, including any prespecified hypotheses | 7 |
| **Methods** | | | |
| Study design | 4 | Present key elements of study design early in the paper | 7 |
| Setting | 5 | Describe the setting, locations, and relevant dates, including periods of recruitment, exposure, follow-up, and data collection | 7-8 |
| Participants | 6 | (*a*) *Cohort study*—Give the eligibility criteria, and the sources and methods of selection of participants. Describe methods of follow-up<br>*Case-control study*—Give the eligibility criteria, and the sources and methods of case ascertainment and control selection. Give the rationale for the choice of cases and controls.<br>*Cross-sectional study*—Give the eligibility criteria, and the sources and methods of selection of participants | 7-8 |
| | | (*b*) *Cohort study*—For matched studies, give matching criteria and number of exposed and unexposed<br>*Case-control study*—For matched studies, give matching criteria and the number of controls per case | NA |
| Variables | 7 | Clearly define all outcomes, exposures, predictors, potential confounders, and effect modifiers. Give diagnostic criteria, if applicable | 8-10 |

| | | | |
|---|---|---|---|
| Data sources/ measurement | 8* | For each variable of interest, give sources of data and details of methods of assessment (measurement). Describe comparability of assessment methods if there is more than one group | 8-10 |
| Bias | 9 | Describe any efforts to address potential sources of bias | 8-11 |
| Study size | 10 | Explain how the study size was arrived at | 8; eFigure 1 |
| Quantitative variables | 11 | Explain how quantitative variables were handled in the analyses. If applicable, describe which groupings were chosen and why | 8-11 |
| Statistical methods | 12 | (*a*) Describe all statistical methods, including those used to control for confounding | 10-12 |
| | | (*b*) Describe any methods used to examine subgroups and interactions | 11-12 |
| | | (*c*) Explain how missing data were addressed | NA |
| | | (*d*) *Cohort study*—If applicable, explain how loss to follow-up was addressed<br>*Case-control study*—If applicable, explain how matching of cases and controls was addressed.<br>*Cross-sectional study*—If applicable, describe analytical methods taking account of sampling strategy | NA |
| | | (*e*) Describe any sensitivity analyses | 11-12 |
| **Results** | | | |
| Participants | 13* | (a) Report numbers of individuals at each stage of study—eg numbers potentially eligible, examined for eligibility, confirmed eligible, included in the study, completing follow-up, and analysed | eFigure 1 |
| | | (b) Give reasons for non-participation at each stage | eFigure 1 |
| | | (c) Consider use of a flow diagram | eFigure 1 |
| Descriptive data | 14* | (a) Give characteristics of study participants (eg demographic, clinical, social) and information on exposures and potential confounders | Table 1 |
| | | (b) Indicate number of participants with missing data for each variable of interest | eFigure 1 |
| | | (c) *Cohort study*—Summarise follow-up time (eg, average and total amount) | 12 |

| | | | |
|---|---|---|---|
| Outcome data | 15* | *Cohort study*—Report numbers of outcome events or summary measures over time | 12 |
| | | *Case-control study*—Report numbers in each exposure category, or summary measures of exposure | NA |
| | | *Cross-sectional study*—Report numbers of outcome events or summary measures | NA |
| Main results | 16 | (*a*) Give unadjusted estimates and, if applicable, confounder-adjusted estimates and their precision (eg, 95% confidence interval). Make clear which confounders were adjusted for and why they were included | 12-14 |
| | | (*b*) Report category boundaries when continuous variables were categorized | N/A |
| | | (*c*) If relevant, consider translating estimates of relative risk into absolute risk for a meaningful time period | 12-13 |
| Other analyses | 17 | Report other analyses done—eg analyses of subgroups and interactions, and sensitivity analyses | 14-15 |
| **Discussion** | | | |
| Key results | 18 | Summarise key results with reference to study objectives | 15 |
| Limitations | 19 | Discuss limitations of the study, taking into account sources of potential bias or imprecision. Discuss both direction and magnitude of any potential bias | 18 |
| Interpretation | 20 | Give a cautious overall interpretation of results considering objectives, limitations, multiplicity of analyses, results from similar studies, and other relevant evidence | 16-18 |
| Generalisability | 21 | Discuss the generalisability (external validity) of the study results | 18 |
| **Other information** | | | |
| Funding | 22 | Give the source of funding and the role of the funders for the present study and, if applicable, for the original study on which the present article is based | 19 |

**eTable 5:** E-values for minimum dose and median VILPA and MV-ILPA values for incident type 2 diabetes

| **VILPA duration (min/day)** | **E-value** |
|---|---|
| Minimum dose (ED50 value) | 1.78 (1.48) |
| Median values | 2.05 (1.63) |
| **VILPA frequency (bouts/day)** | **E-value** |
| Minimum dose (ED50 value) | 1.87 (1.48) |
| Median values | 2.05 (1.58) |
| **MV-ILPA duration (min/day)** | **E-value** |
| Minimum dose (ED50 value) | 1.81 (1.53) |
| Median values | 2.42 (1.86) |
| **MV-ILPA frequency (bouts/day)** | **E-value** |
| Minimum dose (ED50 value) | 1.69 (1.42) |
| Median values | 2.25 (1.70) |

Abbreviations: VILPA, vigorous intermittent lifestyle physical activity; MV-ILPA, moderate-to-vigorous intermittent lifestyle physical activity. E-value represents point estimate (lower limit of the confidence interval) that an unmeasured confounder would need to have with both the exposure and outcome to explain away the exposure-outcome association, conditional on the measured covariates.

**eFigure 1:** Participant selection process

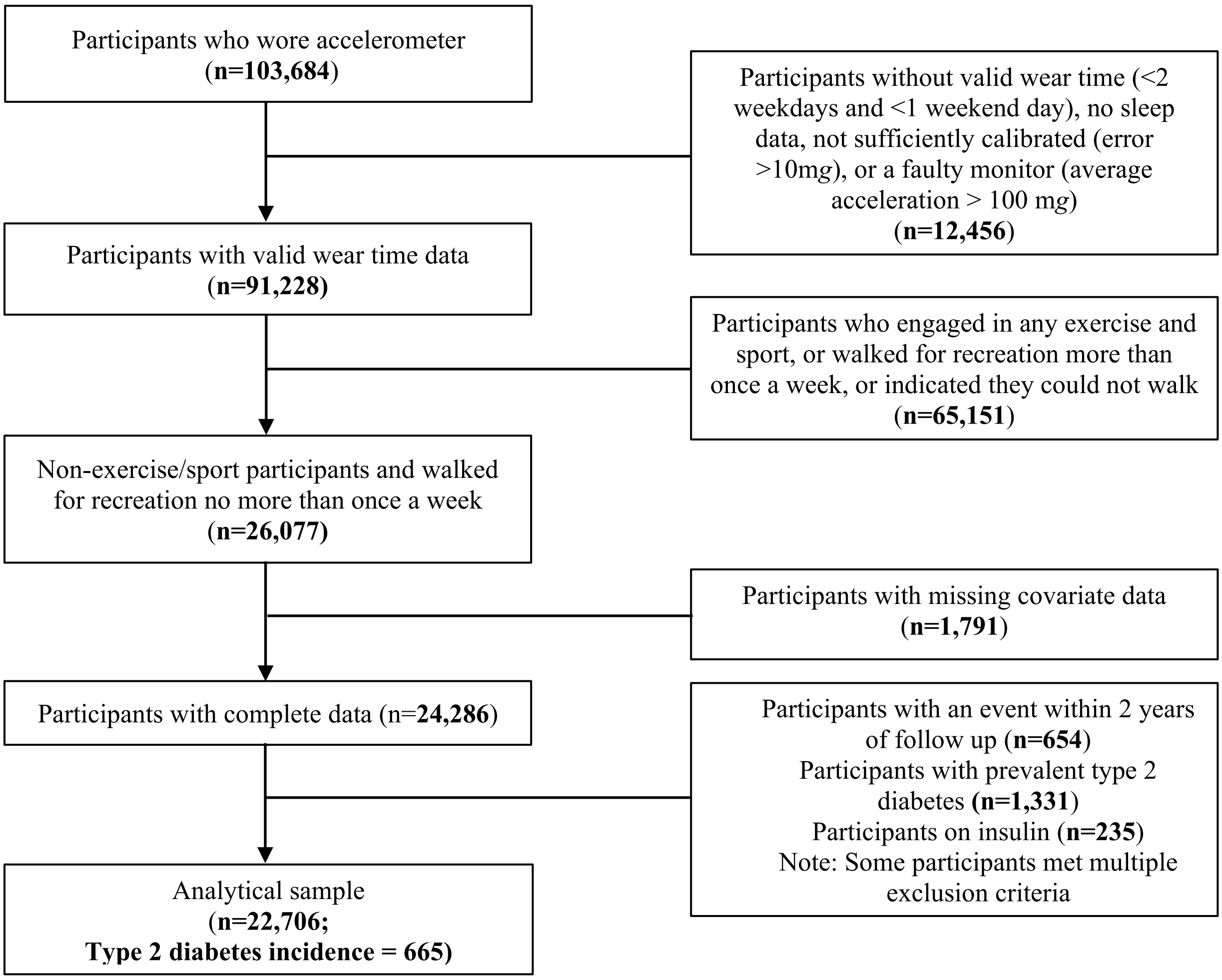

**eFigure 2:** Adjusted absolute risk-based dose response curves of daily VILPA duration and frequency with incident type 2 diabetes (n = 22,706; events = 665)

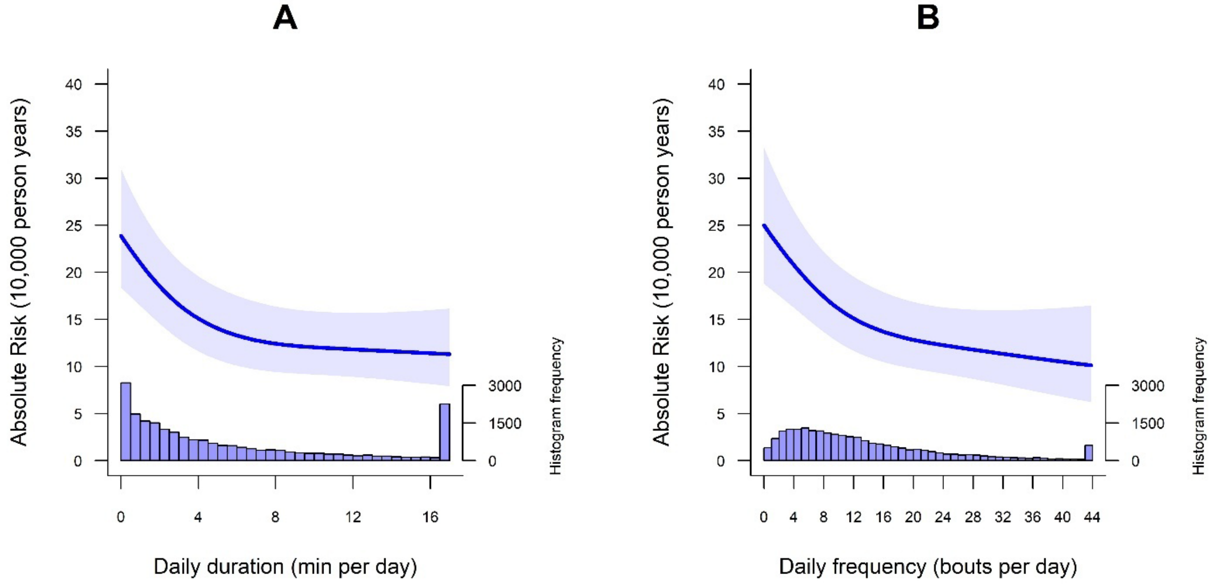


**Legend:** The model was adjusted for sex, age, education levels, ethnicity, fruit and vegetable consumption, smoking status, alcohol consumption, sleep duration, discretionary screen time, medication use (cholesterol and blood pressure lowering), prevalent cancer, prevalent cardiovascular disease (CVD), parental history of cancer, CVD and type 2 diabetes (T2D), and physical activity energy expenditure volume of non-exposure intensity components (light-, moderate- and vigorous-intensity physical activity [excluding VILPA]).

**eFigure 3:** Adjusted absolute risk-based dose response curves of daily MV-ILPA duration and frequency with incident type 2 diabetes (n = 22,706; events = 665)

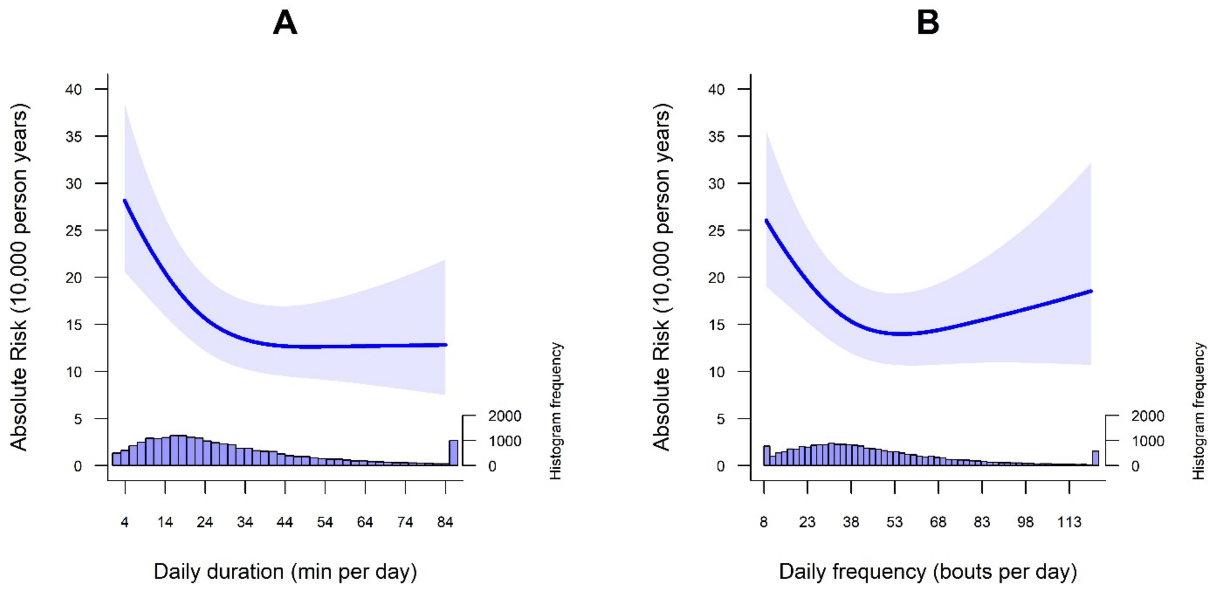


**Legend:** The model was adjusted for sex, age, education levels, ethnicity, fruit and vegetable consumption, smoking status, alcohol consumption, sleep duration, discretionary screen time, medication use (cholesterol and blood pressure lowering), prevalent cancer, prevalent cardiovascular disease (CVD), parental history of cancer, CVD and type 2 diabetes (T2D), and physical activity energy expenditure volume of non-exposure intensity components (light-, moderate- and vigorous-intensity physical activity [excluding MV-ILPA]).

**eFigure 4:** Adjusted dose response curves of daily VILPA duration and frequency with incident type 2 diabetes, with additional adjustment for BMI and other cardiometabolic biomarkers (n = 18,313; events = 526)

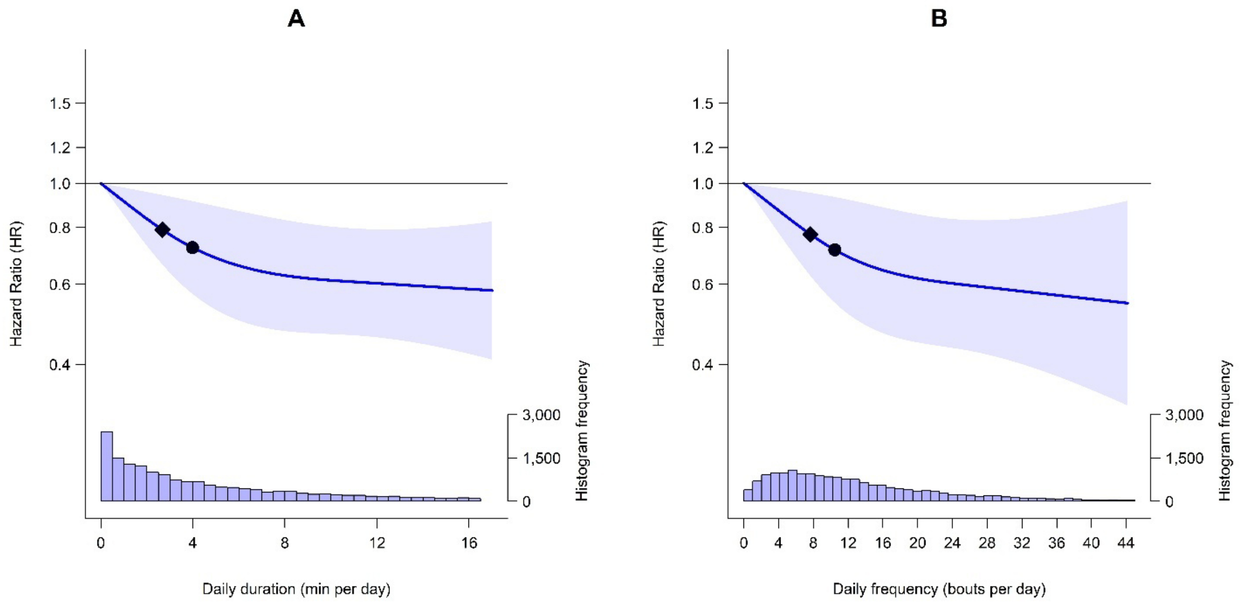


**Legend:** The line represents hazard ratios (HRs) and the shaded area represents their 95% confidence intervals associated with increasing daily (A) duration and (B) frequency of VILPA. The diamond refers to the minimal VILPA duration/frequency dose (as indicated by ED50 statistics) associated with 50% of optimal risk reduction, while the circle refers to the HR associated with the median duration/frequency of VILPA. The histogram on the right shows the sample distribution. The model was adjusted for sex, age, education levels, ethnicity, fruit and vegetable consumption, smoking status, alcohol consumption, sleep duration, discretionary screen time, medication use (cholesterol and blood pressure lowering), prevalent cancer, prevalent cardiovascular disease (CVD), parental history of cancer, CVD and type 2 diabetes (T2D), physical activity energy expenditure volume of non-exposure intensity components (light-, moderate- and vigorous-intensity physical activity [excluding VILPA]), body mass index (BMI) and other cardiometabolic biomarkers (low-density lipoprotein [LDL], high-density lipoprotein [HDL], triglycerides, systolic blood pressure, diastolic blood pressure). The reference point was the minimum data point of VILPA duration (0 min/day) and frequency (0 bouts/day).

**eFigure 5:** Adjusted dose response curves of daily VILPA duration and frequency with incident type 2 diabetes, with additional adjustment for waist circumference and other cardiometabolic biomarkers (n = 18,554; events = 533)

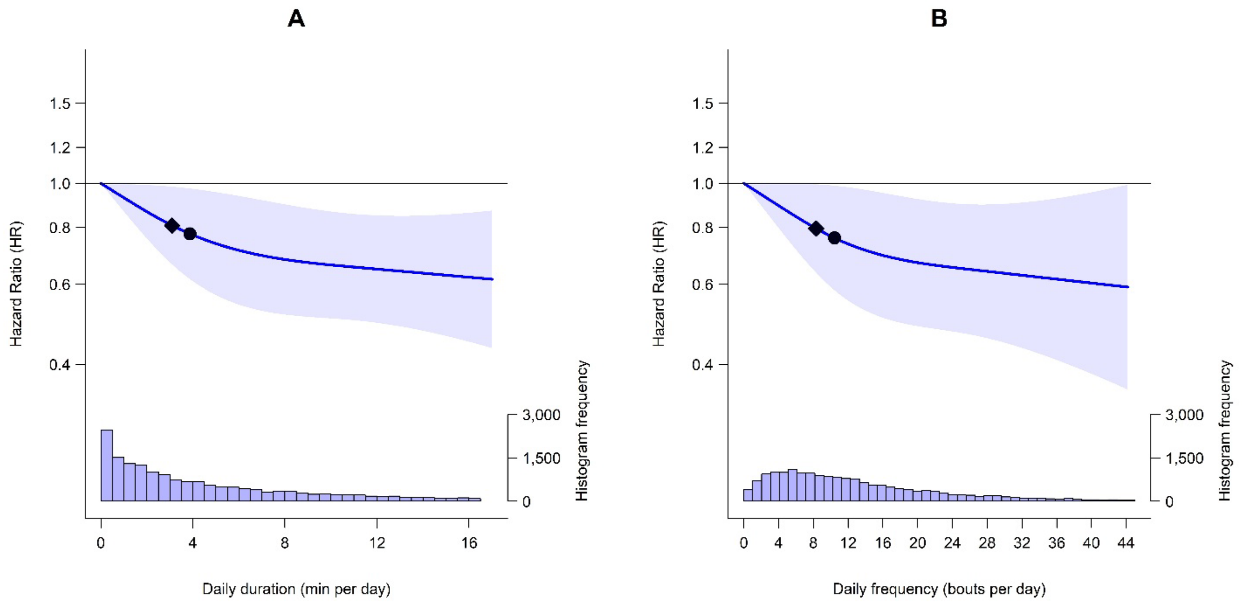


**Legend:** The line represents hazard ratios (HRs) and the shaded area represents their 95% confidence intervals associated with increasing daily (A) duration and (B) frequency of VILPA. The diamond refers to the minimal VILPA duration/frequency dose (as indicated by ED50 statistics) associated with 50% of optimal risk reduction, while the circle refers to the HR associated with the median duration/frequency of VILPA. The histogram on the right shows the sample distribution. The model was adjusted for sex, age, education levels, ethnicity, fruit and vegetable consumption, smoking status, alcohol consumption, sleep duration, discretionary screen time, medication use (cholesterol and blood pressure lowering), prevalent cancer, prevalent cardiovascular disease (CVD), parental history of cancer, CVD and type 2 diabetes (T2D), physical activity energy expenditure volume of non-exposure intensity components (light-, moderate- and vigorous-intensity physical activity [excluding VILPA]), waist circumference and biomarkers (low-density lipoprotein [LDL], high-density lipoprotein [HDL], triglycerides, systolic blood pressure, diastolic blood pressure). The reference point was the minimum data point of VILPA duration (0 min/day) and frequency (0 bouts/day).

**eFigure 6:** Adjusted dose response curves of daily MV-ILPA duration and frequency with incident type 2 diabetes, with additional adjustment for BMI and other cardiometabolic biomarkers (n = 18,313; events = 526)

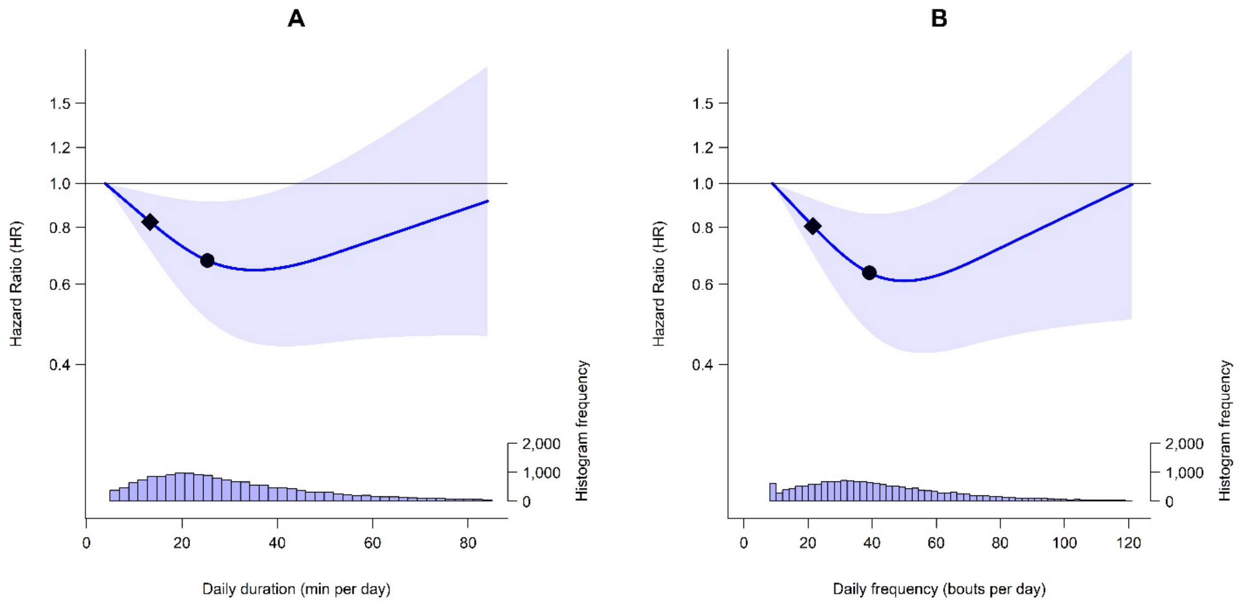


**Legend:** The line represents hazard ratios (HRs) and the shaded area represents their 95% confidence intervals associated with increasing daily (A) duration and (B) frequency of MV-ILPA. The diamond refers to the minimal MV-ILPA duration/frequency dose (as indicated by ED50 statistics) associated with 50% of optimal risk reduction, while the circle refers to the HR associated with the median duration/frequency of MV-ILPA. The histogram on the right shows the sample distribution. The model was adjusted for sex, age, education levels, ethnicity, fruit and vegetable consumption, smoking status, alcohol consumption, sleep duration, discretionary screen time, medication use (cholesterol and blood pressure lowering), prevalent cancer, prevalent cardiovascular disease (CVD), parental history of cancer, CVD and type 2 diabetes (T2D), physical activity energy expenditure volume of non-exposure intensity components (light-, moderate- and vigorous-intensity physical activity [excluding MV-ILPA]), body mass index (BMI) and other cardiometabolic biomarkers (low-density lipoprotein [LDL], high-density lipoprotein [HDL], triglycerides, systolic blood pressure, diastolic blood pressure). The reference point was the minimum data point of MV-ILPA duration (3.9 min/day) and frequency (8.9 bouts/day).

**eFigure 7:** Adjusted dose response curves of daily MV-ILPA duration and frequency with incident type 2 diabetes, with additional adjustment for waist circumference other cardiometabolic biomarkers (n = 18,554; events = 533)

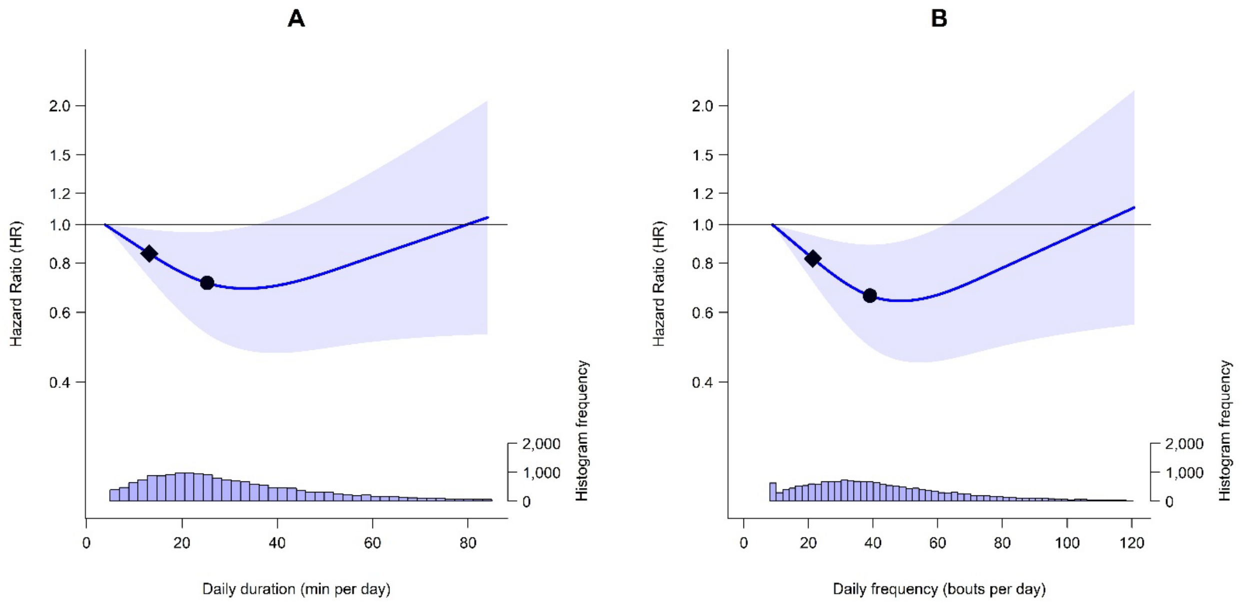


**Legend:** The line represents hazard ratios (HRs) and the shaded area represents their 95% confidence intervals associated with increasing daily (A) duration and (B) frequency of MV-ILPA. The diamond refers to the minimal MV-ILPA duration/frequency dose (as indicated by ED50 statistics) associated with 50% of optimal risk reduction, while the circle refers to the HR associated with the median duration/frequency of MV-ILPA. The histogram on the right shows the sample distribution. The model was adjusted for sex, age, education levels, ethnicity, fruit and vegetable consumption, smoking status, alcohol consumption, sleep duration, discretionary screen time, medication use (cholesterol and blood pressure lowering), prevalent cancer, prevalent cardiovascular disease (CVD), parental history of cancer, CVD and type 2 diabetes (T2D), physical activity energy expenditure volume of non-exposure intensity components (light-, moderate- and vigorous-intensity physical activity [excluding MV-ILPA]), waist circumference and other cardiometabolic biomarkers (low-density lipoprotein [LDL], high-density lipoprotein [HDL], triglycerides, systolic blood pressure, diastolic blood pressure). The reference point was the minimum data point of MV-ILPA duration (3.9 min/day) and frequency (8.9 bouts/day).

**eFigure 8:** Adjusted dose response curves of daily VILPA duration and frequency with incident type 2 diabetes, excluding participants who were underweight (BMI < 18.5 kg/m$^2$) or reported poor health (n = 21,346; events = 584)

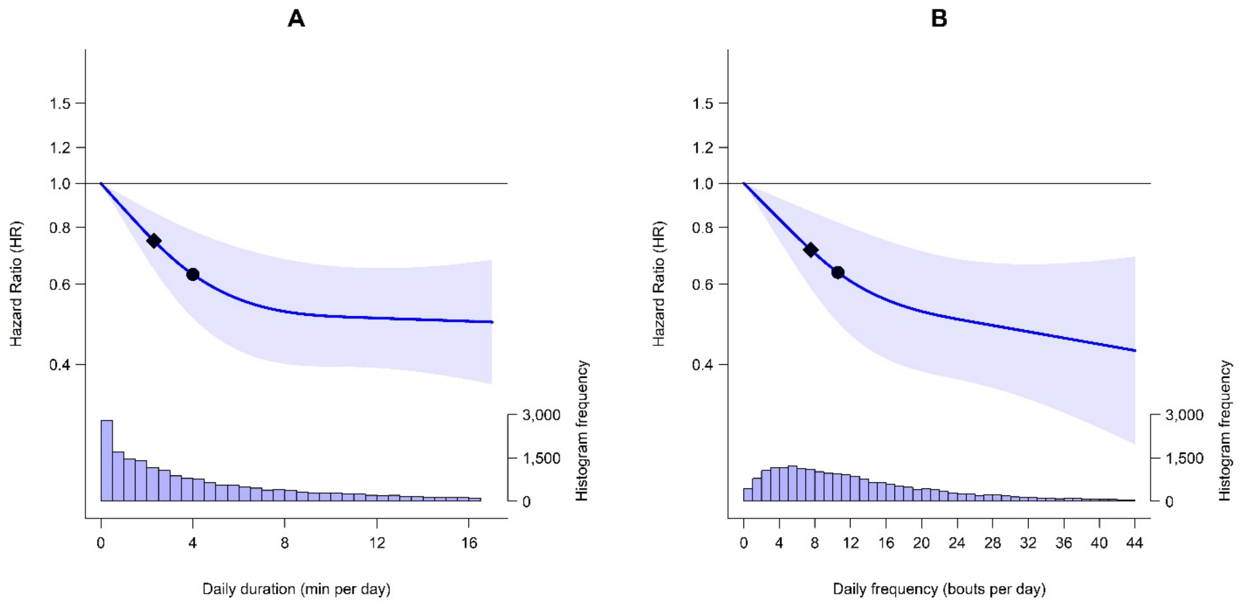


**Legend:** The line represents hazard ratios (HRs) and the shaded area represents their 95% confidence intervals associated with increasing daily (A) duration and (B) frequency of VILPA. The diamond refers to the minimal VILPA duration/frequency dose (as indicated by ED50 statistics) associated with 50% of optimal risk reduction, while the circle refers to the HR associated with the median duration/frequency of VILPA. The histogram on the right shows the sample distribution. The model was adjusted for sex, age, education levels, ethnicity, fruit and vegetable consumption, smoking status, alcohol consumption, sleep duration, discretionary screen time, medication use (cholesterol and blood pressure lowering), prevalent cancer, prevalent cardiovascular disease (CVD), parental history of cancer, CVD and type 2 diabetes (T2D), and physical activity energy expenditure volume of non-exposure intensity components (light-, moderate- and vigorous-intensity physical activity [excluding VILPA]). The reference point was the minimum data point of VILPA duration (0 min/day) and frequency (0 bouts/day).

**eFigure 9:** Adjusted dose response curves of daily VILPA duration and frequency with incident type 2 diabetes, excluding participants with a frailty index score of 3 or higher (on a 0-5 scale) (n = 20,435; events = 570)

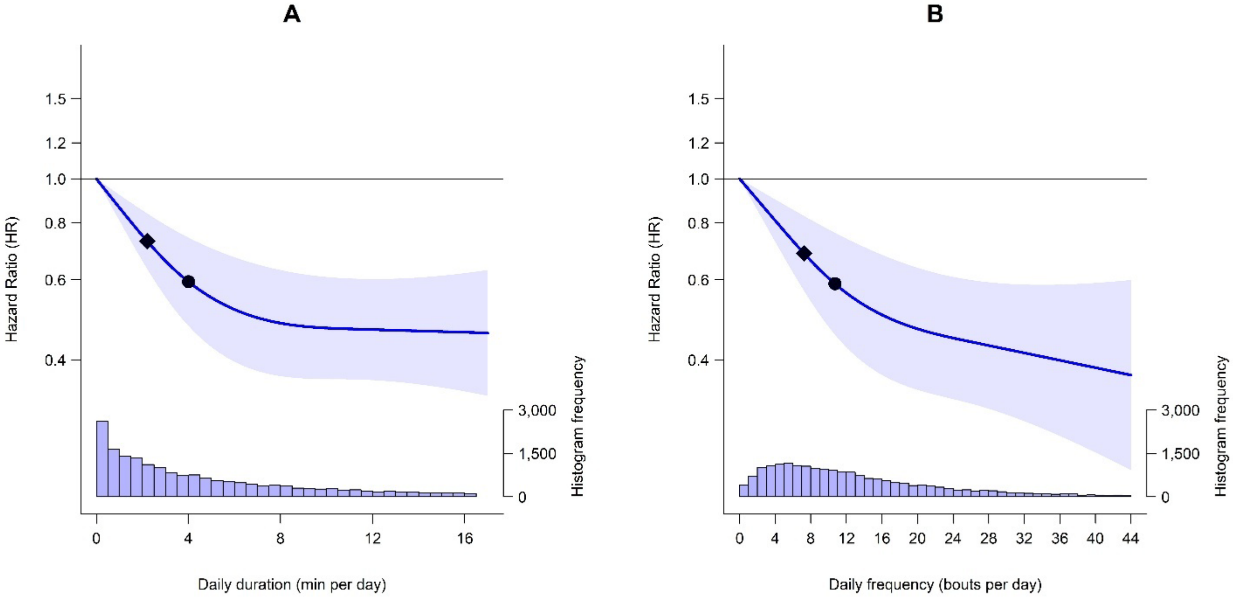


**Legend:** The line represents hazard ratios (HRs) and the shaded area represents their 95% confidence intervals associated with increasing daily (A) duration and (B) frequency of VILPA. The diamond refers to the minimal VILPA duration/frequency dose (as indicated by ED50 statistics) associated with 50% of optimal risk reduction, while the circle refers to the HR associated with the median duration/frequency of VILPA. The histogram on the right shows the sample distribution. The model was adjusted for sex, age, education levels, ethnicity, fruit and vegetable consumption, smoking status, alcohol consumption, sleep duration, discretionary screen time, medication use (cholesterol and blood pressure lowering), prevalent cancer, prevalent cardiovascular disease (CVD), parental history of cancer, CVD and type 2 diabetes (T2D), and physical activity energy expenditure volume of non-exposure intensity components (light-, moderate- and vigorous-intensity physical activity [excluding VILPA]). The reference point was the minimum data point of VILPA duration (0 min/day) and frequency (0 bouts/day).

**eFigure 10:** Adjusted dose response curves of daily MV-ILPA duration and frequency with incident type 2 diabetes, excluding participants who were underweight (BMI < 18.5 kg/m$^2$) or reported poor health (n = 21,346; events = 584)

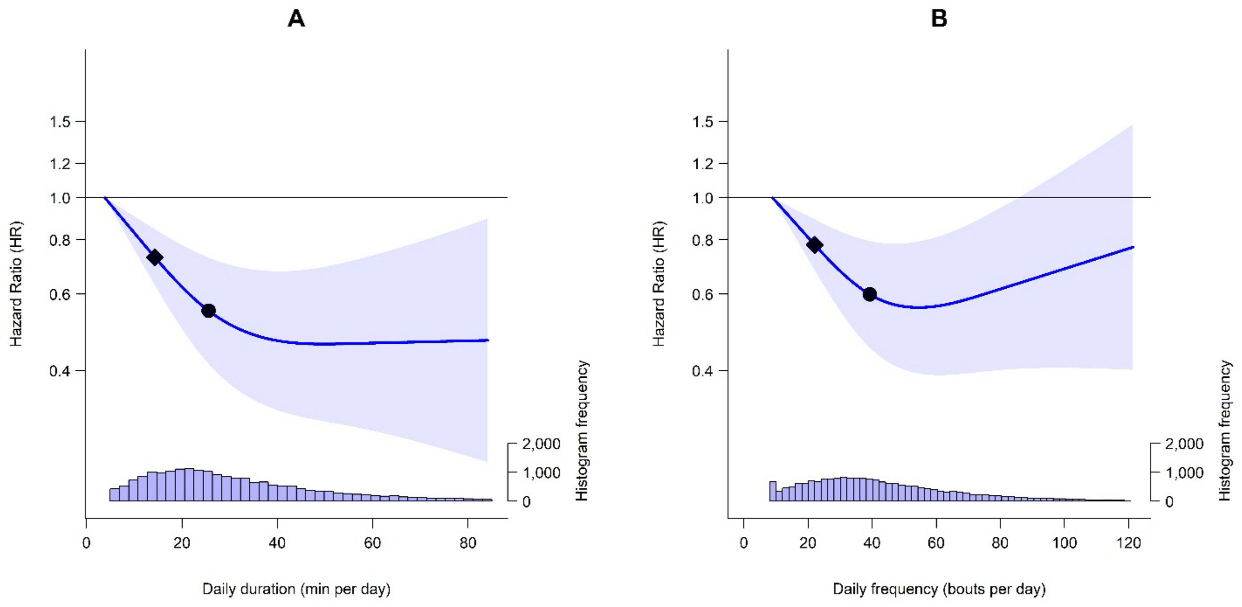


**Legend:** The line represents hazard ratios (HRs) and the shaded area represents their 95% confidence intervals associated with increasing daily (A) duration and (B) frequency of MV-ILPA. The diamond refers to the minimal MV-ILPA duration/frequency dose (as indicated by ED50 statistics) associated with 50% of optimal risk reduction, while the circle refers to the HR associated with the median duration/frequency of MV-ILPA. The histogram on the right shows the sample distribution. The model was adjusted for sex, age, education levels, ethnicity, fruit and vegetable consumption, smoking status, alcohol consumption, sleep duration, discretionary screen time, medication use (cholesterol and blood pressure lowering), prevalent cancer, prevalent cardiovascular disease (CVD), parental history of cancer, CVD and type 2 diabetes (T2D), and physical activity energy expenditure volume of non-exposure intensity components (light-, moderate- and vigorous-intensity physical activity [excluding MV-ILPA]). The reference point was the minimum data point of MV-ILPA duration (3.9 min/day) and frequency (9.0 bouts/day).

**eFigure 11:** Adjusted dose response curves of daily MV-ILPA duration and frequency with incident type 2 diabetes, excluding participants with a frailty index score of 3 or higher (on a 0-5 scale) (n = 20,435; events = 570)

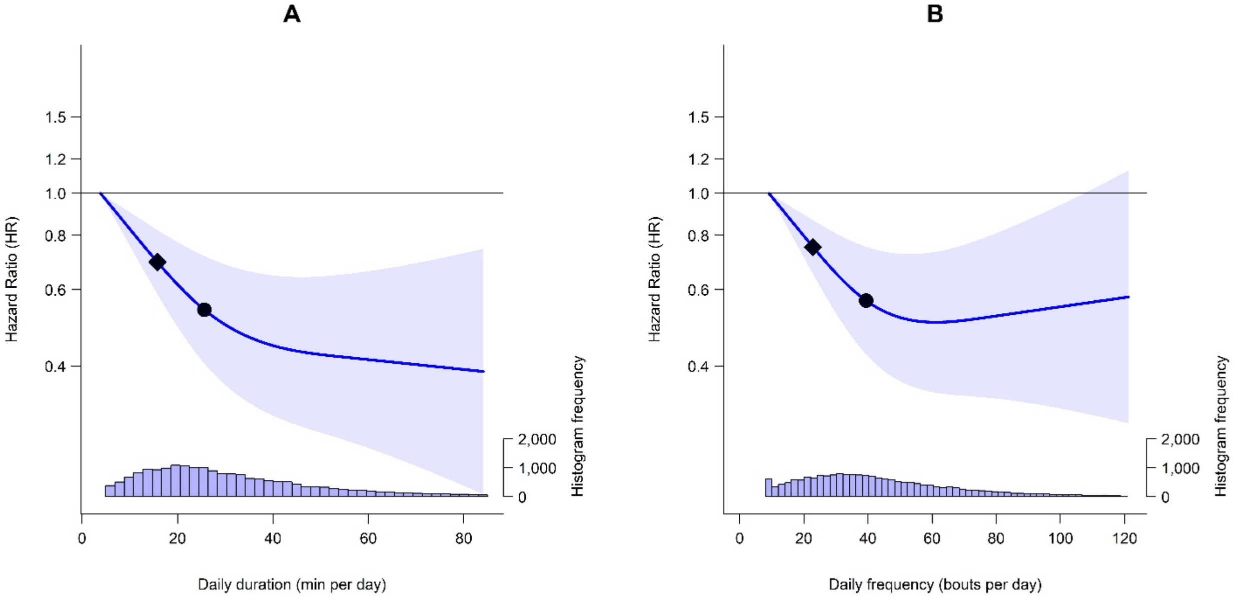


**Legend:** The line represents hazard ratios (HRs) and the shaded area represents their 95% confidence intervals associated with increasing daily (A) duration and (B) frequency of MV-ILPA. The diamond refers to the minimal MV-ILPA duration/frequency dose (as indicated by ED50 statistics) associated with 50% of optimal risk reduction, while the circle refers to the HR associated with the median duration/frequency of MV-ILPA. The histogram on the right shows the sample distribution. The model was adjusted for sex, age, education levels, ethnicity, fruit and vegetable consumption, smoking status, alcohol consumption, sleep duration, discretionary screen time, medication use (cholesterol and blood pressure lowering), prevalent cancer, prevalent cardiovascular disease (CVD), parental history of cancer, CVD and type 2 diabetes (T2D), and physical activity energy expenditure volume of non-exposure intensity components (light-, moderate- and vigorous-intensity physical activity [excluding MV-ILPA]). The reference point was the minimum data point of MV-ILPA duration (3.9 min/day) and frequency (9.1 bouts/day).

**eFigure 12:** Adjusted dose-response curves of daily VILPA duration and frequency with incident type 2 diabetes, excluding participants with prevalent cancer or CVD (n = 18,380; events = 476)

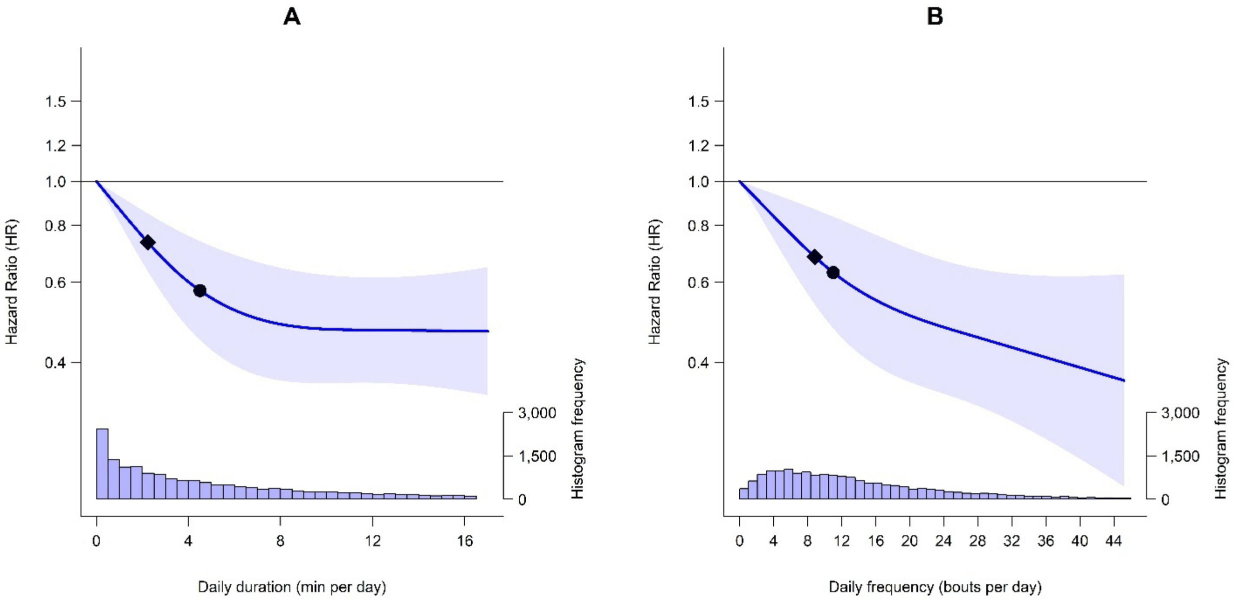


**Legend:** The line represents hazard ratios (HRs) and the shaded area represents their 95% confidence intervals associated with increasing daily (A) duration and (B) frequency of VILPA. The diamond refers to the minimal VILPA duration/frequency dose (as indicated by ED50 statistics) associated with 50% of optimal risk reduction, while the circle refers to the HR associated with the median duration/frequency of VILPA. The histogram on the right shows the sample distribution. The model was adjusted for sex, age, education levels, ethnicity, fruit and vegetable consumption, smoking status, alcohol consumption, sleep duration, discretionary screen time, medication use (cholesterol and blood pressure lowering), parental history of cancer, CVD and type 2 diabetes (T2D), and physical activity energy expenditure volume of non-exposure intensity components (light-, moderate- and vigorous-intensity physical activity [excluding VILPA]). The reference point was the minimum data point of VILPA duration (0 min/day) and frequency (0 bouts/day).

**eFigure 13:** Adjusted dose-response curves of daily MV-ILPA duration and frequency with incident type 2 diabetes, excluding participants with prevalent cancer or CVD (n = 18,380; events = 476)

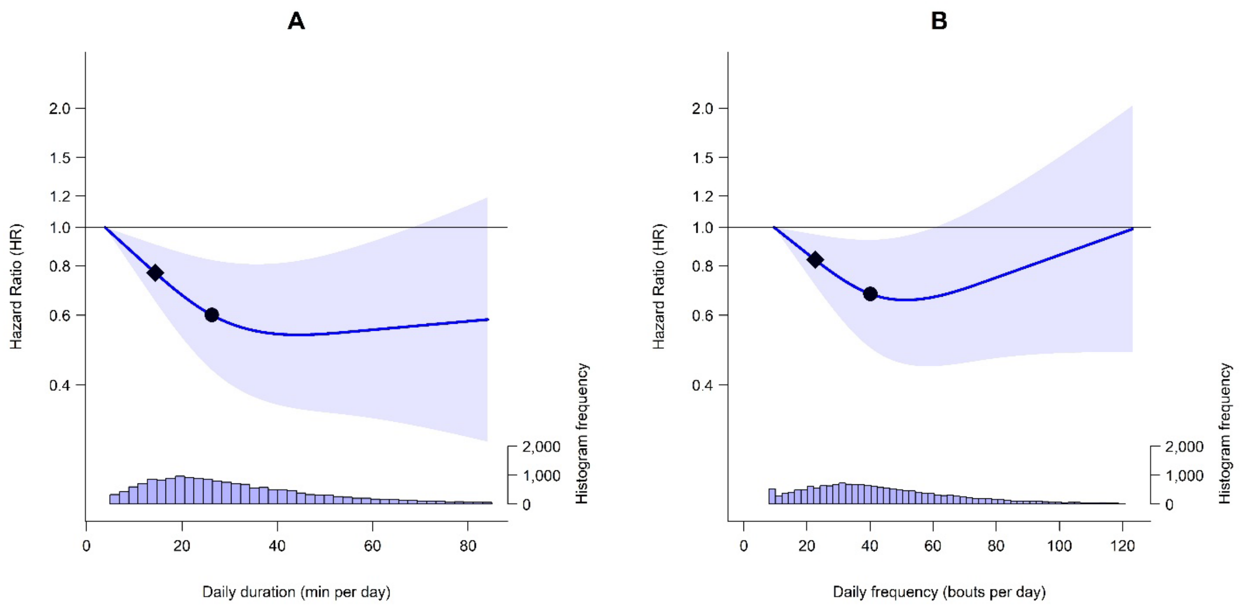


**Legend:** The line represents hazard ratios (HRs) and the shaded area represents their 95% confidence intervals associated with increasing daily (A) duration and (B) frequency of MV-ILPA. The diamond refers to the minimal MV-ILPA duration/frequency dose (as indicated by ED50 statistics) associated with 50% of optimal risk reduction, while the circle refers to the HR associated with the median duration/frequency of MV-ILPA. The histogram on the right shows the sample distribution. The model was adjusted for sex, age, education levels, ethnicity, fruit and vegetable consumption, smoking status, alcohol consumption, sleep duration, discretionary screen time, medication use (cholesterol and blood pressure lowering), parental history of cancer, CVD and type 2 diabetes (T2D), and physical activity energy expenditure volume of non-exposure intensity components (light-, moderate- and vigorous-intensity physical activity [excluding MV-ILPA]). The reference point was the minimum data point of MV-ILPA duration (3.9 min/day) and frequency (9.6 bouts/day).

**eFigure 14:** Adjusted dose response curves of daily VILPA duration and frequency with incident type 2 diabetes using the 25th percentile of data point as the reference (n = 22,706; events = 665)

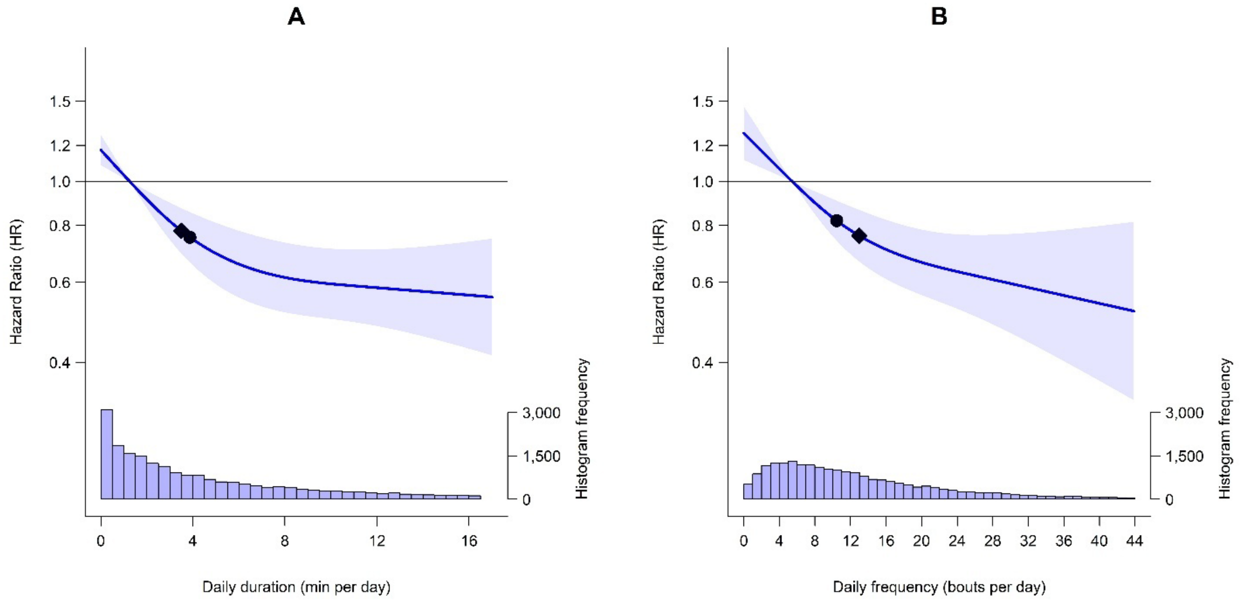


**Legend:** The line represents hazard ratios (HRs) and the shaded area represents their 95% confidence intervals associated with increasing daily (A) duration and (B) frequency of VILPA. The diamond refers to the minimal VILPA duration/frequency dose (as indicated by ED50 statistics) associated with 50% of optimal risk reduction, while the circle refers to the HR associated with the median duration/frequency of VILPA. The histogram on the right shows the sample distribution. The model was adjusted for sex, age, education levels, ethnicity, fruit and vegetable consumption, smoking status, alcohol consumption, sleep duration, discretionary screen time, medication use (cholesterol and blood pressure lowering), prevalent cancer, prevalent cardiovascular disease (CVD), parental history of cancer, CVD and type 2 diabetes (T2D), and physical activity energy expenditure volume of non-exposure intensity components (light-, moderate- and vigorous-intensity physical activity [excluding VILPA]). The reference point was the 25th percentile of VILPA duration (1.3 min/day) and bouts (5.4 bouts/day) distribution.

**eFigure 15:** Adjusted dose response curves of daily MV-ILPA duration and frequency with incident type 2 diabetes using the 25th percentile of data point as the reference point (n = 22,706; events = 665)

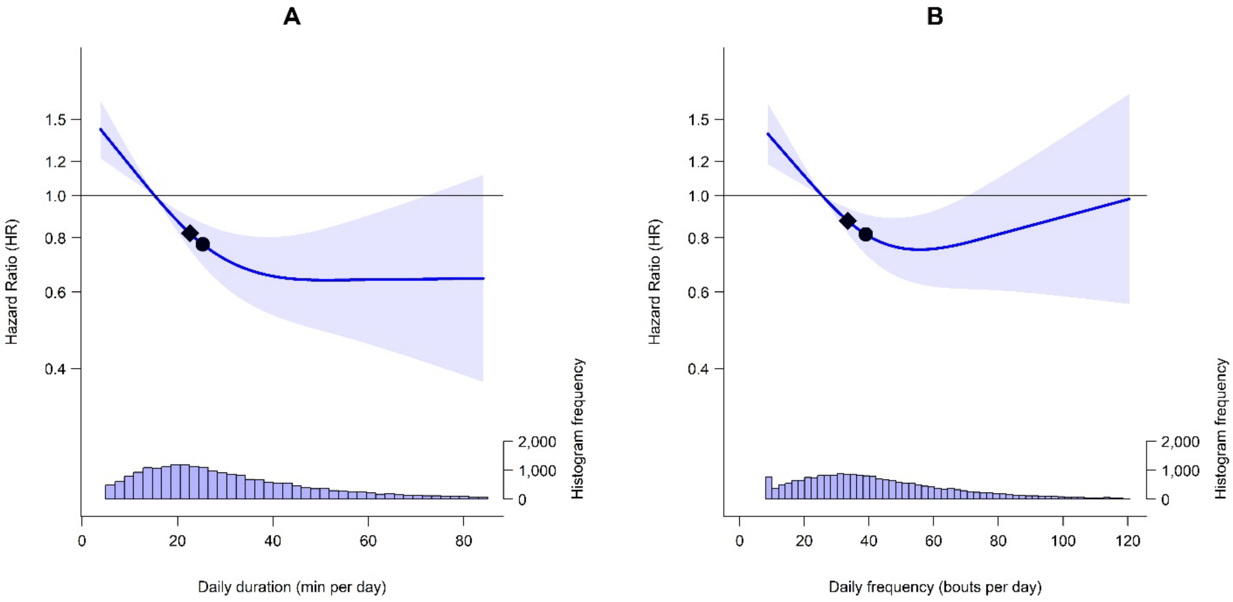


**Legend:** The line represents hazard ratios (HRs) and the shaded area represents their 95% confidence intervals associated with increasing daily (A) duration and (B) frequency of MV-ILPA. The diamond refers to the minimal MV-ILPA duration/frequency dose (as indicated by ED50 statistics) associated with 50% of optimal risk reduction, while the circle refers to the HR associated with the median duration/frequency of MV-ILPA. The histogram on the right shows the sample distribution. The model was adjusted for sex, age, education levels, ethnicity, fruit and vegetable consumption, smoking status, alcohol consumption, sleep duration, discretionary screen time, medication use (cholesterol and blood pressure lowering), prevalent cancer, prevalent cardiovascular disease (CVD), parental history of cancer, CVD and type 2 diabetes (T2D) and physical activity energy expenditure volume of non-exposure intensity components (light-, moderate- and vigorous-intensity physical activity [excluding MV-ILPA]). The reference point was set at 25th percentile of daily MV-ILPA duration (15.3 min/day) and frequency (25.6 bouts/day) distribution.

**eFigure 16:** Adjusted dose response curves of daily VILPA duration and frequency with incident type 2 diabetes using alternative knots placement (10th, 33rd and 67th percentiles) (n = 22,706; events = 665)

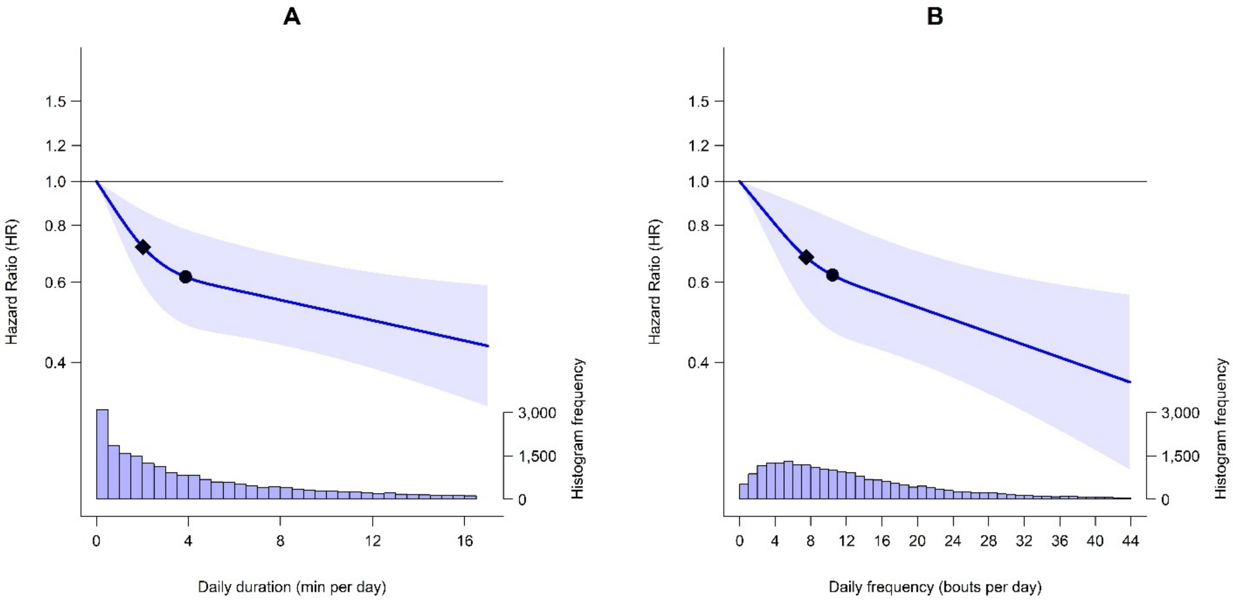


**Legend:** The line represents hazard ratios (HRs) and the shaded area represents their 95% confidence intervals associated with increasing daily (A) duration and (B) frequency of VILPA. The diamond refers to the minimal VILPA duration/frequency dose (as indicated by ED50 statistics) associated with 50% of optimal risk reduction, while the circle refers to the HR associated with the median duration/frequency of VILPA. The histogram on the right shows the sample distribution. The model was adjusted for sex, age, education levels, ethnicity, fruit and vegetable consumption, smoking history, alcohol consumption, sleep duration, discretionary screen time, medication use (cholesterol and blood pressure lowering), prevalent cancer, prevalent cardiovascular disease (CVD), family history of cancer, CVD and type 2 diabetes (T2D), and physical activity energy expenditure volume of non-exposure intensity components (light-, moderate- and vigorous-intensity physical activity [excluding VILPA]). The reference point was the minimum data point of VILPA duration (0 min/day) and frequency (0 bouts/day).

**eFigure 17:** Adjusted dose response curves of daily MV-ILPA duration and frequency with incident type 2 diabetes using alternative knots placement (10th, 33rd and 67th percentiles) (n = 22,706; events = 665)

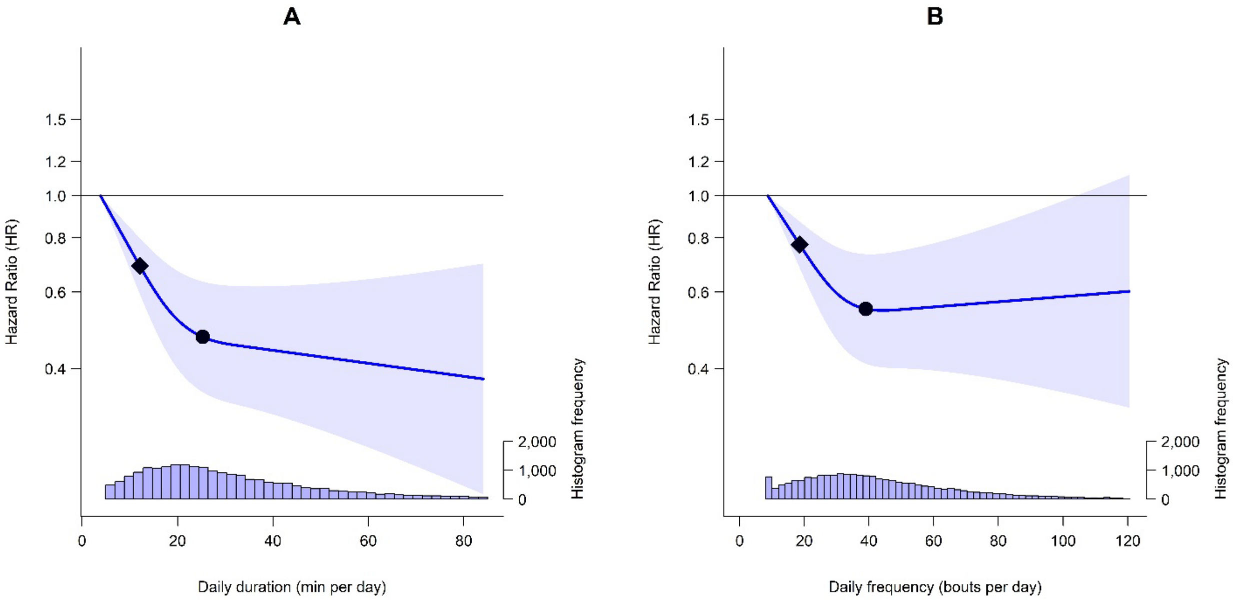


**Legend:** The line represents hazard ratios (HRs) and the shaded area represents their 95% confidence intervals associated with increasing daily (A) duration and (B) frequency of MV-ILPA. The diamond refers to the minimal MV-ILPA duration/frequency dose (as indicated by ED50 statistics) associated with 50% of optimal risk reduction, while the circle refers to the HR associated with the median duration/frequency of MV-ILPA. The histogram on the right shows the sample distribution. The model was adjusted for sex, age, education levels, ethnicity, fruit and vegetable consumption, smoking status, alcohol consumption, sleep duration, discretionary screen time, medication use (cholesterol and blood pressure lowering), prevalent cancer, prevalent cardiovascular disease (CVD), parental history of cancer, CVD and type 2 diabetes (T2D) and physical activity energy expenditure volume of non-exposure intensity components (light-, moderate- and vigorous-intensity physical activity [excluding MV-ILPA]). The reference point was the minimum data point of MV-ILPA duration (3.9 min/day) and frequency (8.7 bouts/day).

**eFigure 18:** Sex-specific adjusted dose response curves of daily VILPA duration and frequency with incident type 2 diabetes (male: n = 9,908; events = 400; female, n = 12,798; events = 265)

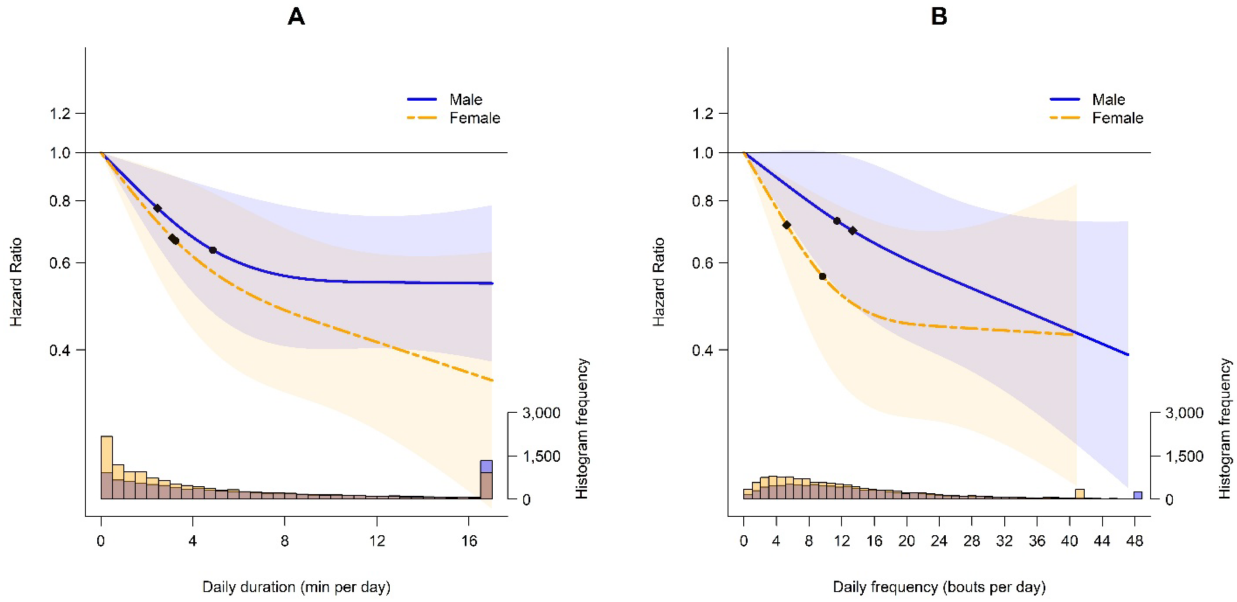


**Legend:** The line represents hazard ratios (HRs) and the shaded area represents their 95% confidence intervals associated with increasing daily (A) duration and (B) frequency of VILPA. The diamond refers to the minimal VILPA duration/frequency dose (as indicated by ED50 statistics) associated with 50% of optimal risk reduction, while the circle refers to the HR associated with the median duration/frequency of VILPA. The histogram on the right shows the sample distribution. The model was adjusted for age, education levels, ethnicity, fruit and vegetable consumption, smoking status, alcohol consumption, sleep duration, discretionary screen time, medication use (cholesterol and blood pressure lowering), prevalent cancer, prevalent cardiovascular disease (CVD), parental history of cancer, CVD and type 2 diabetes (T2D), and physical activity energy expenditure volume of non-exposure intensity components (light-, moderate- and vigorous-intensity physical activity [excluding VILPA]). The reference point was the minimum data point of VILPA duration (0 min/day for both males and females) and frequency (0 bouts/day for both males and females).

**eFigure 19:** Sex-specific adjusted dose response curves of daily MV-ILPA duration and frequency with incident type 2 diabetes (male: n = 9,908; events = 400; female, n = 12,798; events = 265)

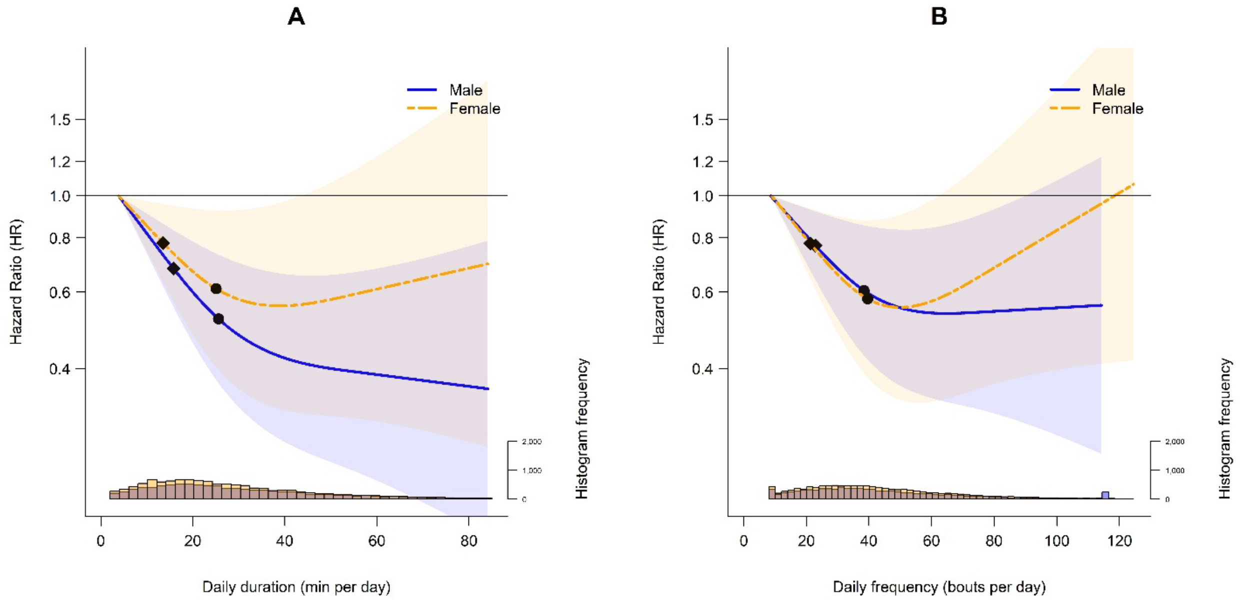


**Legend:** The line represents hazard ratios (HRs) and the shaded area represents their 95% confidence intervals associated with increasing daily (A) duration and (B) frequency of MV-ILPA. The diamond refers to the minimal MV-ILPA duration/frequency dose (as indicated by ED50 statistics) associated with 50% of optimal risk reduction, while the circle refers to the HR associated with the median duration/frequency of MV-ILPA. The histogram on the right shows the sample distribution. The model was adjusted for age, education levels, ethnicity, fruit and vegetable consumption, smoking status, alcohol consumption, sleep duration, discretionary screen time, medication use (cholesterol and blood pressure lowering), prevalent cancer, prevalent cardiovascular disease (CVD), parental history of cancer, CVD and type 2 diabetes (T2D) and physical activity energy expenditure volume of non-exposure intensity components (light-, moderate- and vigorous-intensity physical activity [excluding MV-ILPA]). The reference point was the minimum data point of MV-ILPA duration (3.9 min/day for both males and females) and frequency (8.6 bouts/day for males and 8.9 bouts/day for females).

**eMethod:** Physical activity classification

**Wearable device-based physical activity classification**

**eMethod – Figure 1** below summarises how physical activity intensity was classified using a previously validated wrist-worn accelerometery Random Forest (RF) activity classifier.[2] RF is an ensemble of multiple decision trees. Each tree is learned on a bootstrap sample of training data and each node in the tree is split using the best among a randomly selected set of acceleration features. The decisions from each tree are aggregated and a final model prediction is based on majority vote. The RF model requires very little pre-processing of the data, as the features do not need to be normalised. Additionally, the model is resistant to over fitting the training data because each tree within the forest is independently grown to maximum depth using a randomly selected subset of features.

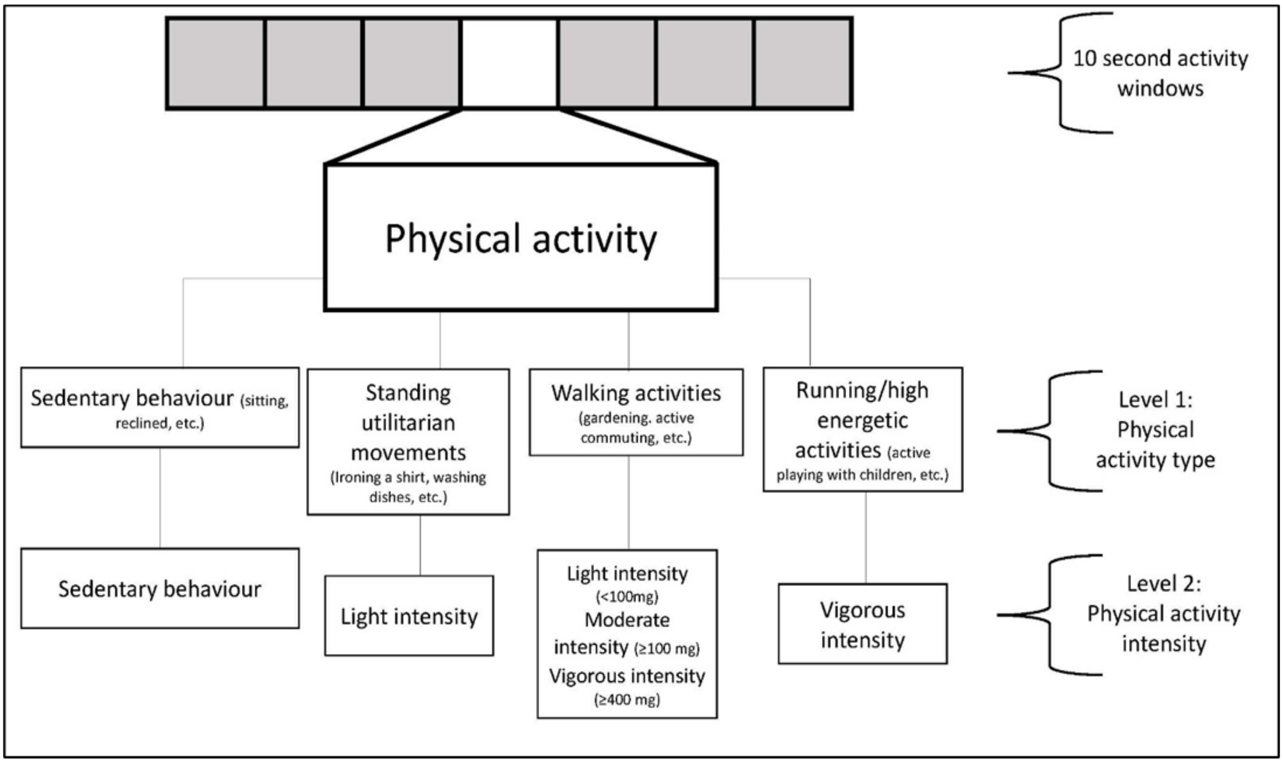


**eMethod – Figure 1:** Physical activity type and intensity diagram describing the 2-level (stage) intensity classification schema

This 2-stage classifier which first categorised physical activity in 10 second windows into one of four activity classes: sedentary, standing utilitarian movements (e.g., ironing a shirt, washing dishes), walking (e.g., gardening, active commuting, mopping floors), and running/high energetic activities (e.g., active playing with children). These activity classes were then assigned to one of four activity intensities: sedentary, light, moderate, and vigorous. Walking activities were classified as light (an acceleration value of < 100 m*g*), moderate (≥ 100 m*g*), and vigorous (≥ 400 m*g*) intensity. For example, for a VPA bout to last for 1.5 minutes (90 seconds), nine consecutive 10-second windows needed to be classified as vigorous. Differentiation from sleep[3] and non-wear[4] was identified using the change in tilt angle and acceleration standard deviation.

Monitors were calibrated[5] and corrected for orientation[6] using previously published methods although residual signal and alignment uncertainties may persist.

**Physical activity classification performance**

The performance of this physical activity classification scheme was tested in an independent sample of 102 participants from the US (University of California Irvine Center for Machine Learning and Intelligent Systems - *Physical Activity Monitoring for Aging People* study [published data], accessible at https://archive.ics.uci.edu/ml/datasets)[7] and Australia (University of Queensland *Where and When at Work* study [published data], and University of Sydney *Intermittent Lifestyle Physical Activity* Study [unpublished data]).[8] This data includes direct observation measurement of 103,607 activity samples from structured and free-living activities (17,267 minutes), which were used to assess the robustness and generalisability of the classifier (**eMethod – Table 1**). For free-living activities, participant-worn or researcher-held Go-Pro video-recordings were used to attain ground-truth physical activity. Video files were imported into the Noldus Observer XT software version 16.0 for continuous direct observation coding. A two-stage direct observation scheme was implemented in which the participant's movement behaviour was coded for activity type and then activity intensity based on Compendium of Physical Activities.[9] The direct observation system generated a vector of date-time stamps corresponding to the start and finish of each movement event, which were used to assign the activity codes to the corresponding time segments of the accelerometer data. Interobserver reliability was assessed by dual coding. The intraclass correlation coefficient for coding activities was 0.91 (0.87-0.94).

**eMethod – Table 1:** Intensity classification performance in 102 US and Australian adults (exercisers and non-exercisers pooled; age = 55.8 ± 12.4; 55.8% female) providing 105,767 activity samples from structured (exercised-based) and free-living activities (17,627 minutes) (from published and unpublished data)

| | Sensitivity | Specificity | Precision | F-score | Overall Accuracy | Weighted Kappa | Overall F-score |
|---|---|---|---|---|---|---|---|
| Sedentary | 86.5 | 93.7 | 90.5 | 88.5 | | | |
| Light | 71.2 | 89.4 | 55.8 | 62.6 | | | |
| Moderate | 85.4 | 96.6 | 92.7 | 88.9 | | | |
| Vigorous | 95.4 | 99.4 | 94.6 | 95.0 | | | |
| | | | | | 84.6 | 0.78 | 83.8 |

Performance was further evaluated in a sample of 151 adults (age range 18-91 years, 65.6% female; Method – Figure 2) recruited from the UK (University of Oxford *Capture 24* study [published data], that is publicly accessible at https://ora.ox.ac.uk/objects/uuid:99d7c092-d865-4a19-b096-cc16440cd001).[10] Participants in this study wore body cameras that provided pictures every 20 seconds to annotate ground-truth free-living activity labels. The picture-based activity coding scheme has been previously described.[10] A total of 172,360 activity samples (28,727 minutes) were provided by participants.

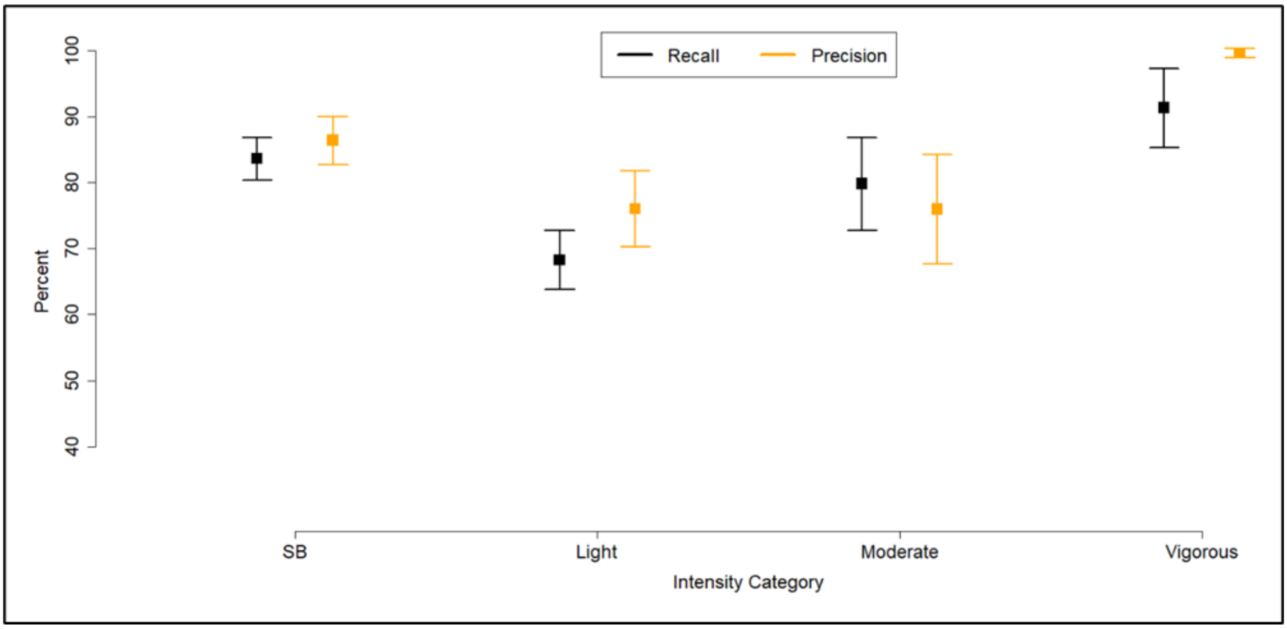


**eMethod – Figure 2:** Participant-level specific recall and precision of activity classification in 151 UK adults

**Confusion Matrix for Activity Classification in US and Australian Adults**

| | Sedentary | Light | Moderate | Vigorous |
|---|---|---|---|---|
| Sedentary | **36,904** | 5,232 | 508 | 2 |
| Light | 3,120 | **11,712** | 1,612 | 17 |
| Moderate | 502 | 4,016 | **29,528** | 526 |
| Vigorous | 226 | 17 | 214 | **9,470** |

Rows = ground truth; columns = predictions; bold = correct labels; numbers represent each 10-second window; all activities were free-living or simulated free-living activities.

From a subset of the participants who had both ground-truth physical activity type-specific data ascertained through video recordings and the physical activity compendium, and physical activity intensity using indirect calorimetry (both gold-standard measures), we assessed intensity classification for individual activity types presented in the "expanded confusion matrix" below. Because of the >100 specific types of physical activity recorded we present the 3 most prevalent activity types for each intensity category.

**Expanded confusion matrix of most prominent activity types within each intensity band in the independent validity testing (n=91; 245,945 seconds of activity data)**

| | | Sedentary | Light | Moderate | Vigorous |
|---|---|---|---|---|---|
| Sedentary | Desk/computer work | **90.2%** | 9.8% | - | - |
| | Sitting/lying | **93.9%** | 6.1% | - | - |
| | Sitting using phone/appliance | **89.4%** | 10.6% | - | - |

| | | | | | |
|---|---|---|---|---|---|
| Light | Washing dishes/kitchen activities | 17.3% | **80.8%** | 2.9% | - |
| | Household chores standing | 12.5% | **83.6%** | 3.9% | - |
| | Slow walking/grocery shopping | 15.2% | **77.8%** | 7.0% | - |
| Moderate | Walking briskly/fast (e.g. for transportation) | - | 11.4% | **86.2%** | 2.4% |
| | Household chores cleaning rooms/ambulation | - | 12.2% | **87.6%** | 0.2% |
| | Occupation brisk walking carrying light objects | - | 11.8% | **87.5%** | 0.7% |
| Vigorous | Manual work | - | 3.0% | 1.3% | **95.7%** |
| | Very fast walking/burst of running (e.g for transportation) | - | 0.8% | 2.3% | **96.9%** |
| | Heavy household outdoor chores | - | 2.4 | 4.2% | **93.4%** |

Rows = ground truth; columns = predictions; bold = correct classification.

To evaluate the capacity of the classifier specifically in the context of incidental physical activity, we also provide a confusion matrix in eMethod - Table 2 below showing the performance among participants who self-reported as non-exercisers in the independent validity testing (n=88; 3688 minutes of video-coded activity data).

**eMethod – Table 2:** Confusion matrix of incidental physical activity among participants who self-identified as non-exercisers in the independent validity testing (n=82; 3688 minutes of activity data)

| | Sedentary | Light | Moderate | Vigorous |
|---|---|---|---|---|
| Sedentary | **92.4%** | 7.6% | - | - |
| Light | 13.3% | **80.8%** | 5.9% | - |
| Moderate | - | 11.7% | **88.1%** | 0.2% |
| Vigorous | - | 1.2% | 1.5% | **97.3%** |

Rows = ground truth; columns = predictions; bold = correct classification; all activities were free-living or simulated free-living activities
